# PBPK Model-Based Prediction of Optimal Rosuvastatin Dosing with Concomitant Ticagrelor in Elderly Patients Considering ABCG2 Genetic Variant

**DOI:** 10.64898/2026.10.16.26365646

**Authors:** Hoang Dat Nguyen, Vinh Hoa Pham, Junjie Ding, Joel Tarning

## Abstract

**Aims:** Rosuvastatin is commonly co-administered with ticagrelor in patients with acute coronary syndrome. Ticagrelor inhibits breast cancer resistance protein (BCRP/ABCG2), increasing rosuvastatin exposure and potentially the risk of rhabdomyolysis. This risk may be further influenced by age and the ABCG2 c.421C>A variant. We aimed to predict optimal rosuvastatin dosing with ticagrelor in elderly patients stratified by ABCG2 genotype using physiologically based pharmacokinetic (PBPK) modelling

**Methods:** PBPK models for rosuvastatin and ticagrelor were developed and validated in PK-Sim® using clinical data from healthy adults, elderly patients, and individuals with wild-type or homozygous ABCG2 c.421C>A genotypes. Key parameters, including BCRP catalytic activity and age-dependent declines in CYP2C9 and CYP3A4 activity, were optimized. The drug–drug interaction model incorporated competitive BCRP inhibition by ticagrelor. Rosuvastatin steady-state exposure was simulated with half or quarter standard doses during ticagrelor co-administration, and exposure matching approach was used to identify optimal dosing.

**Results:** The PBPK models demonstrated good predictive performance across populations and genotypes. In young adults with the wild-type genotype, halving the rosuvastatin dose resulted in an AUC fold change of 1.44, supporting a half-dose strategy. In young homozygous variant carriers, half-dose co-administration produced comparable exposure (AUC fold change 0.78). In elderly wild-type individuals, a four-fold dose reduction achieved comparable exposure (AUC fold change 1.06), whereas elderly variant carriers required only a half-dose reduction (1.12).

**Conclusions:** PBPK modelling supported age- and ABCG2 genotype-specific rosuvastatin dosing with ticagrelor. Elderly wild-type patients may require a further dose reduction to minimise toxicity while maintaining therapeutic exposure.

## 1. Introduction

Rosuvastatin is a 3-hydroxy-3-methylglutaryl coenzyme A inhibitors that commonly used in the treatment of hypercholesterolemia[1]. Compared to other statin drug, rosuvastatin is more effective for reducing low-density lipoprotein cholesterol[2]. Rosuvastatin has a low oral bioavailability of 20% [3] and excreted mainly unchanged into urine and bile[1], with approximately 10% of drug metabolized by cytochrome P450 (CYP) 2C9 and CYP2C19[4]. Rosuvastatin is a substrate of transporter OATP2B1 and BCRP. Following absorption into the systemic circulation, its uptake into hepatocytes is driven largely by OATP1B1 and OATP1B3, with NTCP and OATP2B1 playing a minor role. Hepatic elimination occurs predominantly through biliary secretion, in which BCRP is the principal efflux pathway, and with a lesser extent by MRP2[5]. Additionally, genetic variant of BCRP is an important factor associated with rosuvastatin PK exposure. ABCG2 rs2231142 (c.421C>A) associates with a decreased function of ABCG2[6]. The homozygous variant of ABCG2 rs2231142 associates with a 2 folds increase in PK exposure compared to wildtype[7]. The frequency of this variant is substantial across different populations, with reported variant allele frequencies of 0.11, 0.19 and 0.20 in European-American, Mexican-American and American Indian population, respectively[8], while as it ranged 0.076-0.21 in Asian population[9].

Ticagrelor, a carbocyclic nucleoside analogue, acts as a reversible and selective inhibitor of the platelet P2Y12 receptor. It is absorbed quickly with a Tmax of 1.3–3 h, and is primarily metabolized through CYP3A4 to form an equipotent active metabolite, AR-C124910XX[10]. For acute coronary syndrome, the recommended first-line therapy for the first year is dual antiplatelet treatment with aspirin plus a P2Y12 inhibitor (ticagrelor or prasugrel). Strict LDL cholesterol control is also recommended to lower the risk of recurrent atherothrombotic events[11]. Therefore, ticagrelor is frequently prescribed with rosuvastatin in clinical practice.

Rhabdomyolysis is a very rare but severe adverse event for statins treatment. This adverse event occurs in a dose-depend manner and estimated to be 4 per 10,000 patients using high-dose statin and 1 per 10,000 patients receiving standard statin dosing[12]. A > 2-fold increase in PK exposure of rosuvastatin co-medicated with ticagrelor has been reported, driven by ticagrelor-mediated inhibition of ABCG2[1]. This higher PK exposure potentially increases the risk of rhabdomyolysis[13]. Moreover, the presence of ACCG2 variant may further complicate the magnitude of this DDI effect.

In addition, age is an important factor influencing plasma exposure of ticagrelor. It has been reported that AUC and Cmax increased 52% and 63% in elderly compared to the young adults respectively[14]. Age-related physiological changes, including alterations in body composition, organ mass, and hepatic blood flow—particularly reduced liver size and perfusion—may lead to increased bioavailability and reduced clearance of ticagrelor. Furthermore, an age-associated decline in hepatic CYP3A activity could further contribute to slower metabolism and higher systemic exposure[14]. Therefore, Therefore, advanced age may further exacerbate ticagrelor-mediated increases in rosuvastatin exposure, particularly in elderly patients carrying ABCG2 variants. Currently, no specific dose recommendation is available for rosuvastatin when administered concomitantly with ticagrelor, particularly in elderly patients carrying ABCG2 variants.

In the context of DDI involving combined factors of age and ABCG2, physiologically based pharmacokinetic (PBPK) modeling, a mechanistic *in silico* approach, offers a powerful tool to quantitatively evaluate this complex drug-drug-gene-age interaction[15]. This study aimed to use a PBPK modeling approach to predict the optimal rosuvastatin dose during its interaction with ticagrelor, considering both ABCG2 wild-type and homozygous variant genotypes in elderly populations.

## 2. Method

### 2.1 Software

The PBPK model for ticagrelor and rosuvastatin was developed using the PK-Sim® software (version 12.0). Visualization of PBPK results and PK exposure calculations were performed using R (version 4.0.2, The R Foundation for Statistical Computing) within RStudio (version 4.2.3, RStudio, Inc., Boston, MA, USA).

### 2.2 PBPK Modeling workflow

The model development and validation process adhered to established best practices for PBPK modeling, as outlined in relevant guidelines[16].

The PBPK modeling workflow for the ticagrelor and rosuvastatin DDI is shown in Fig 1. In brief, rosuvastatin and ticagrelor PBPK models will be developed and validated in various population regarding age (non-elderly and elderly participants) and ABCG2 genotypes, respectively. A PBPK model for rosuvastatin was initially optimized and verified using data from non-elderly adults. The verified model was subsequently extrapolated to and validated in an elderly population. The model was then expanded to characterize PK across various ABCG2 c.421C>A genotypes. Using a similar stepwise strategy, a PBPK model for ticagrelor was first optimized and verified in non-elderly subjects and subsequently extrapolated and validated in the elderly subjects.

**Fig 1:**
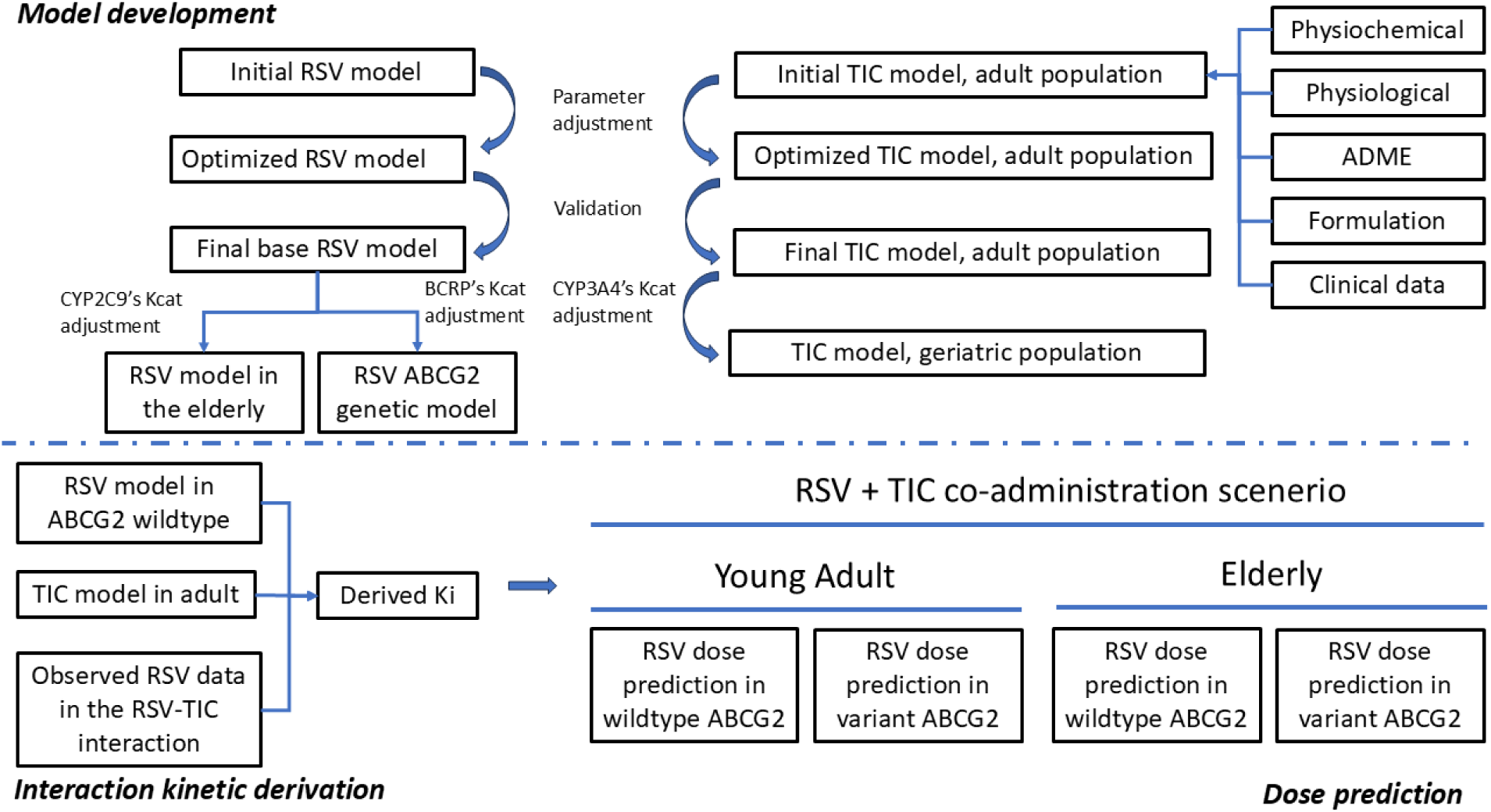
Workflow for PBPK model development, validation, and clinical application. *Footnote:* RSV, rosuvastatin; TIC, ticagrelor. Models were developed and validated for RSV (base model in adult, geriatric and *ABCG2* c.421C>A wild-type, and variant) and TIC (base) in adult and geriatric populations. The final models were applied to predict RSV dosing during DDI with TIC across genotypes and age groups

**Fig 2:**
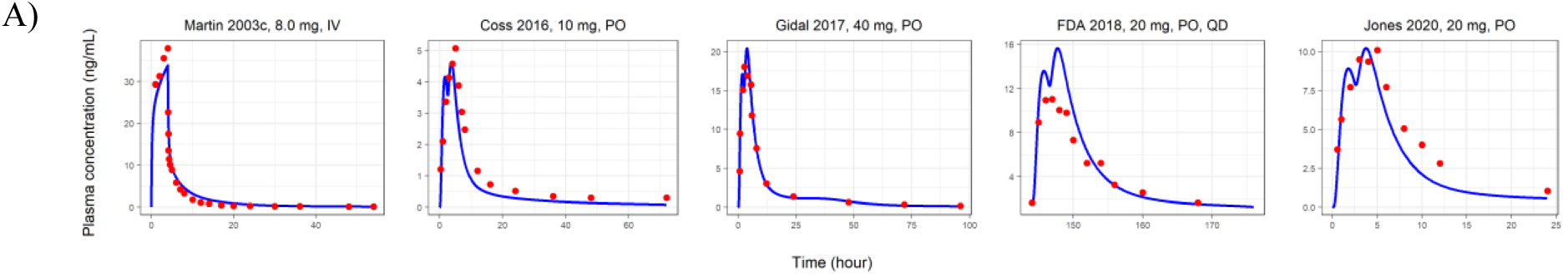
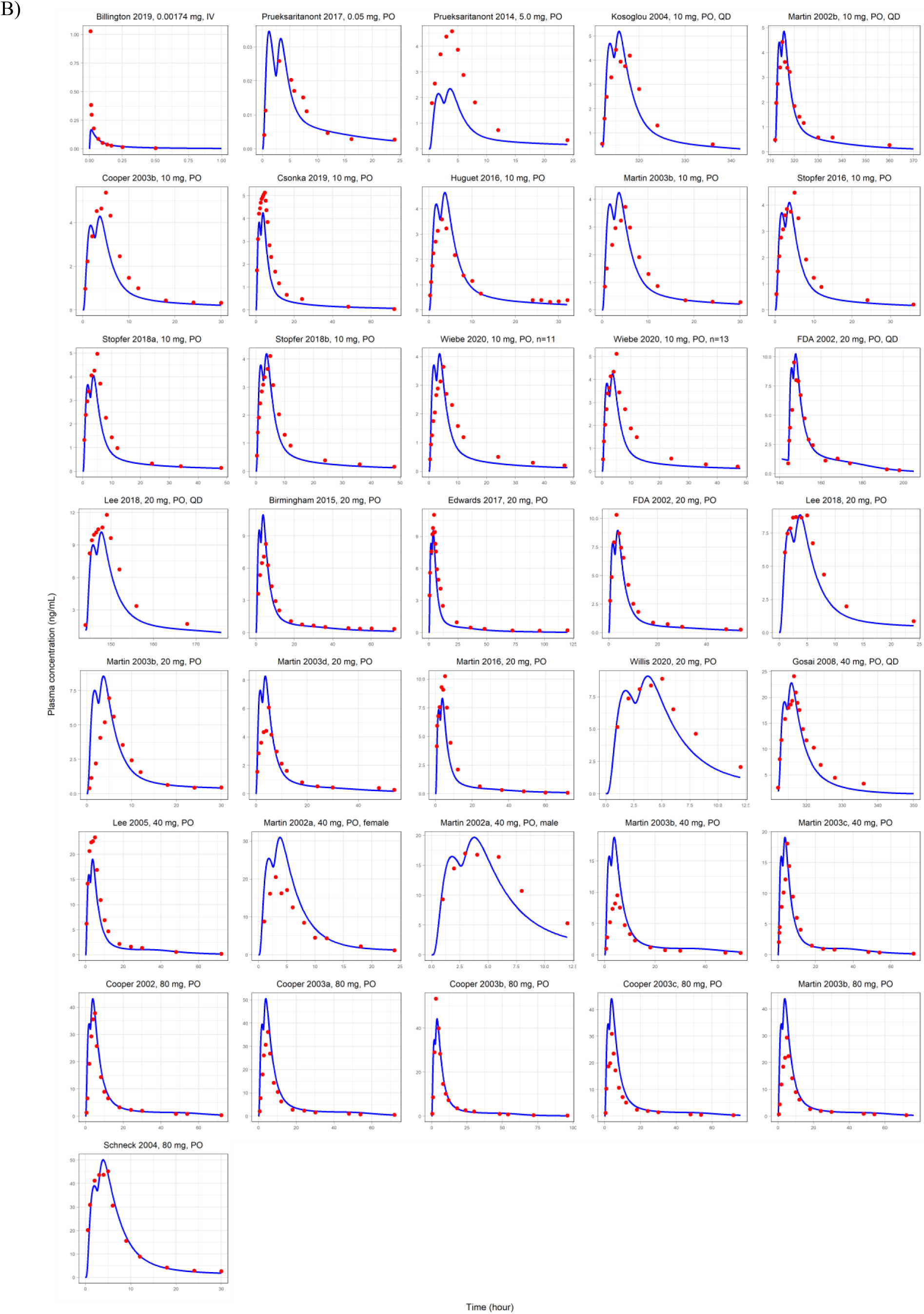
Rosuvastatin plasma concentration–time profiles in A) training dataset, B) validation data set of base models. Simulations are shown as lines, observed data are shown as dots. Details on administration protocols, study populations, and literature references are listed in previous study (1)

Sequentially, the DDI model between rosuvastatin and ticagrelor was developed and verified. The DDI model was then applied to simulate the rosuvastatin PK during co-administration with ticagrelor in elderly patients stratify by ABCG2 genotypes.

#### 2.2.1 Rosuvastatin PBPK model

In order to assess BCRP impact on PK expsoure in the previous published PBPK model [17], a sensitivity analysis was conducted and revealed that a 100-fold reduction in BCRP activity (Kcat from 1.01 min⁻¹ to 0.01 min⁻¹) resulted in only a modest (∼10%) increase in rosuvastatin Cmax at a 10 mg dose. This minimal predicted change contrasts the 131% increase in Cmax reported clinically for individuals homozygous for the ABCG2 c.421C>A variant (low BCRP activity) compared to wild-type controls[7], suggesting that the impact of this genotype was not fully captured by modulating BCRP activity alone.

First, intravenous PK data were used to optimize model parameters governing the distribution phase (cellular permeability) and elimination phase (Kcat of BCRP, P-gp, and CYP2C9; Km of BCRP). Initial parameterization assuming equal contribution of P-gp and BCRP Kcat (19 and 18 /min, respectively), resulted in a significant underprediction. To address this, BCRP Km was optimized from 2.02 µmol/L to approximately 17 µmol/L to align with its increased Kcat. Sequentially, CYP2C9 Kcat and permeability parameters were further optimized to retain model performance. The verified IV PBPK model was then extended to oral administration by refining absorption-related parameters, including intestinal permeability.

Second, the model was scaled to the elderly virtual population via scaling of anthropometric, anatomical, and physiological parameters, as implemented in PK-Sim. In addition, the activity of CYP2C9 in elderly population has been reported to be approximately 40% lower than the young adult[18]. Therefore, the Kcat of CYP2C9 was reduced by 40% in the elderly model. The activity of other enzymes and transporters including BCRP were kept unchanged as there is no clear and consistent evidence that the activity of these proteins changes with age.

To further assess the suitability of optimized kinetic parameters related to P-gp and BCRP, the rosuvstatin PBPK model, including genetic model for ABCG wildtype, was further evaluated in a DDI scenario with rifampin which acts as an inhibitor and inducer of P-gp and inhibitor of BCRP. During this step, the Kcat and Km values for the ABCG2 c.421C>A genotype were further optimized and verified.

#### 2.2.2 Ticagrelor PBPK model

A ticagrelor intravenous PBPK model was first developed and verified in HVs population. Key drug-specific parameters for PBPK model were initially identified from the literature[19, 20]. The partition coefficient, the organ permeability and CYP kcat were optimized to best describe the PK data after IV dosing. For the oral model, the intestinal permeability was further optimized against oral PK data.

Second, the verified PBPK model was extended to an elderly virtual population (> 65 years), by integrating age dependent whole-body anatomical and physiological parameters[21]. The aged dependent reduction of CYP3A4 activity was described using the following equation 1 [22]

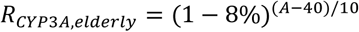

Where RCYP_3A, elderly_ is the activity of CYP3A4 in elderly population, A is age in years.

#### 2.2.3 DDI model

The interaction between ticagrelor and rosuvastatin mediated through ticagrelor inhibition of ABCG2, was modelled by incorporating an inhibition constant (Ki). The Ki parameter was further refined by observed rosuvastatin PK data in DDI clinical studies.

The inhibition of BCRP and P-gp was described using a competitive inhibition model, as defined by the equations below:

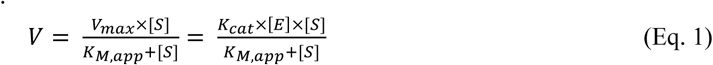

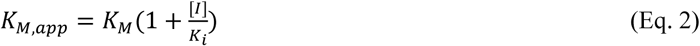

In this model, V is the reaction velocity, Vmax is the maximum reaction velocity, [S] is the free substrate concentration, K_M,app_ is the Michaelis-Menten constant in the presence of inhibitor, Kcat is the catalytic rate constant, [E] is the transporter concentration, K_M_ is the Michaelis-Menten constant in the absence of inhibitor, [I] is the free inhibitor concentration, and Ki is the dissociation constant of the inhibitor-enzyme complex.

The rifampin-mediated induction of P-gp was described using an Emax model, as described in equations 3 and 4.

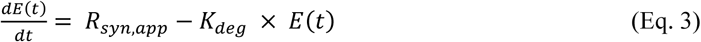

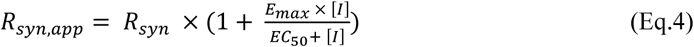

where dE(t)/dt is transporter turnover, R_syn,app_ denotes a zero order rate of transporter synthesis in the presence of an inducer, K_deg_ is the first order degradation rate constant, E(t) is the transporter concentration, R_syn_ is the rate of enzyme or transporter synthesis in the absence of inducer, E_max_ is the maximum induction effect in vivo, [I] is the free inducer concentration, and EC_50_ is the rifampin concentration associated with producing half of the maximum effect in vivo.

### 2.3 Clinical PK data

#### 2.3.1 Rosuvastatin

A previously rosuvastatin PBPK model has been developed and validated using a general Caucasian population without explicit consideration of ABCG2 c.421C>A polymorphism. In the present work, we aimed to explicitly characterize PK for elderly population and subjects with wild-type and homozygous ABCG2 c.421C>A genotypes, the additional published PK data by ABCG2 genotypes were further systematically collected.

For the general population (without specifying ABCG2 genotype), 41 of the 42 study arms utilized in our previous work were retained. One study (Wu et al., 2017) reporting rosuvastatin PK specifically in individuals with the wild-type ABCG2 c.421C>A genotype was used for genotype-specific analysis. A single clinical study was identified to support model validation for geriatric population. For the evaluation of ABCG2 c.421C>A variants carriers, the analysis incorporated five study arms for wild-type subjects and one study arm for homozygous. Detail of clinical studies use for rosuvastatin model development and validation in elderly population and in wildtype and homozygous genotype of ABCG2 c.421C>A variants groups were shown in table S5.

#### 2.3.2 Ticagrelor

A systematic search of the PubMed database was conducted using the keywords “ticagrelor AND pharmacokinetics” between January 2010 and July 2025. Eligible studies included those with PK profiles and parameters under fasted conditions (following single or multiple doses) together with clinical information, including drug dose and demographic data (age, weight, and gender). Studies that were inaccessible or published in languages other than English were excluded. In addition, the present study focuses on Caucasian population because rosuvastatin PBPK model was developed and validated for Caucasian population.

In total, 7 clinical PK studies comprising 25 distinct study arms (23 arms in young adult and 2 arms in elderly population), were included for model development and evaluation. The dose range across these studies varied from 15 to 600 mg, administered as single or multiple doses. Three selected arms from 3 studies (including an IV arm) were used as a training data set, while the remaining study arms were used as the validation data set (Fig 1). Detail of clinical studies used for development and validation of ticagrelor model in young adult and geriatric population were shown in table S10.

A clinical DDI study reporting the interaction between ticagrelor and rosuvastatin has been previously published[1]. This study was used to simulate the drug interaction between ticagrelor and rosuvastatin and derived the inhibition constant, Ki, of ticagrelor toward BCRP.

### 2.4 Simulations

The PBPK DDI model was developed and verified using a built-in PK-SIM virtual population, with the key characteristics (such as, race, sex, age and weight) being aligned with that reported in the clinical PK study. If the range of age and weight was not reported, individual simulations were conducted instead of population simulations.

Human physiological parameters were derived from the default database in PK-Sim®. Relative tissue expression levels of enzymes and transporters were obtained from reverse transcription polymerase chain reaction data available in the PK-Sim® database. The dosage regimen (including administration route, dose amount, frequency) in the simulation was identical to that used in the clinical PK studies.

### 2.5 Model Evaluation

The performance of the PBPK model was assessed by comparing the predicted and observed plasma concentration-time profiles, fraction of drug excreted unchanged in urine (fe), and the two main drug exposure parameters in plasma (AUC and Cmax).

The mean relative deviation (MRD) was calculated to evaluate the predictive performance of plasma drug concentrations (Eq. 5), while the geometric mean fold error (GMFE) was calculated to assess the predictive performance in PK exposures (AUC and Cmax) (Eq. 6).

Additionally, AUC and Cmax ratio were also calculated (Eq.7 and 8). MRD and GMFE values ≤ 2 indicate acceptable model performance. Two-fold error was considered as acceptance criteria for AUC and Cmax predictions. A stricter 1.5-fold error was also summarized.

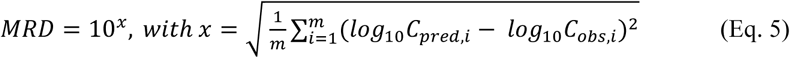

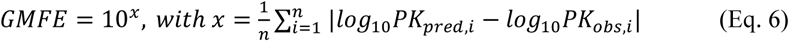

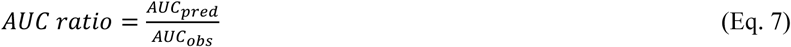

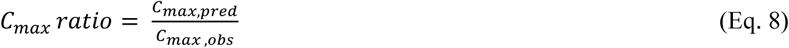

where C_pred,i_ is the predicted plasma concentration, C_obs,i_ is the observed plasma concentration, m is the number of observed concentrations, PK_pred,i_ is the predicted PK parameter value (fe, AUC or Cmax), PK_obs,i_ is the observed PK parameter value, n is the number of studies.

The predictive performance for DDI assessment was conducted by comparing simulated and observed plasma exposure of the victim drug in the presence of ticagrelor [20]. For each DDI study, the ratio of exposure in presence versus absence of ticagrelor was calculated for both observed and simulated data. Model accuracy across studies was summarized using the geometric mean fold error (GMFE), calculated for the predicted DDI AUC and Cmax ratios as shown in Eq. 9 and 10. The GMFE within a twofold range was considered acceptable.

In addition, prediction success was assessed using the criteria proposed by Guest et al. (2011)[23], which applies variable acceptance limits. This approach narrows the acceptance range for weak interactions (ratios near 1) and widens it for stronger interactions, preventing false credit for poor predictions at low potency.

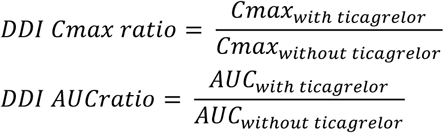

### 2.6 Prediction of optimal dose of rosuvastatin in the interaction with ticagrelor

An exposure-matching approach was used to determine the optimal rosuvastatin dose during co-administration with ticagrelor, with the objective of achieving systemic exposure comparable to rosuvastatin monotherapy.

The final PBPK model was employed to simulate rosuvastatin exposure across four clinically relevant virtual populations: (i) adults with the ABCG2 c.421C>A wild-type genotype, (ii) adults with the homozygous variant genotype, (iii) elderly individuals (ages 65–85 years) with the wild-type genotype, and (iv) elderly individuals with the homozygous variant genotype. Each scenario included 100 virtual subjects, with ages uniformly distributed.

The clinical simulation scenarios included both rosuvastatin monotherapy and co-administration with ticagrelor. In monotherapy, subjects were assumed to receive a rosuvastatin once daily (QD) for two weeks at clinical relevant doses of 5, 10, 20, or 40 mg [24] and the PBPK predicted steady state AUC was considered as the reference value. In the DDI scenarios, ticagrelor was administered twice daily (BID) with a 180 mg loading dose, followed by 90 mg maintenance doses for two weeks, rosuvastatin was administered at the full dose or reduced doses (half-dose or quarter-dose). The PBPK predicted rosuvastatin AUC was then compared to the monotherapy reference value. The dose achieving a comparable AUC (ideally within 0.80 – 1.25 bioequivalence range), was considered the optimal dose.

## 3 Result

### 3.1 Rosuvastatin PBPK model

#### 3.1.1 Base model optimization and validation

The IV PBPK model for rosuvastatin was successfully developed through calibration of key physiological parameters. Distribution parameters, specifically cellular permeability, and elimination parameters—including Kcat values for P-gp and BCRP-mediated transport, along with the Km for BCRP—were optimized to enhance predictive accuracy. Sequentially, an oral PBPK model was developed and validated.

The final optimized PBPK model parameters for rosuvastatin are summarized in Table S1. Model performance is illustrated in Figs2A and 2B, which show good agreement between predicted and observed plasma profiles in the training and validation datasets, respectively. In addition, FigS1 further demonstrates the model’s predictive capability for fraction excreted in urine.

The proposed PBPK model showed good correlation between predicted and observed AUC, Cmax, plasma concentration across both training and validation dataset, as depicted in Figs2B, 3B, 4B S2A, S3A and S4A, respectively. The MRD for plasma concentration was 1.59 (range: 1.24–2.24), with 36 of 41 study arms exhibiting an MRD < 2 as shown in table S2. Overall, the GMFE for AUC was 1.31 (range: 1.0–1.7) and for Cmax was 1.24 (range: 1.0–1.7), indicating robust predictive performance and reliability across multiple validation metrics (Table S3).

#### 3.1.2 DDI model with rifampin

The DDI model with rifampin indicated a good predictive performance, the predicted rosuvastatin concentration profile well captured the observed concentration-time profile, as shown in figS5, while that of fraction of rosuvastatin excreted in urine described in figS6. Overall, the magnitude of DDI effect was adequately predicted as shown in FigS7 and Table S4. The model accurately reproduced the observed DDI effects, with predicted-to observed DDI AUC ratios < 1.4-fold and DDI Cmax ratios < 1.8-fold. Except for a single study arm, all simulated DDI ratios satisfied the prediction success criteria proposed by Guest et al. The outlier case corresponded to the Cmax DDI ratio reported by Prueksaritanont et al. (2014) following co-administration of 5 mg rosuvastatin with 600 mg oral rifampin, for which a predicted-to-observed DDI AUC ratio of 1.8 was obtained. Although this result slightly exceeded acceptance threshold, it remained within the commonly applied two-fold error margin. The overall GMFE was 1.29 (range: 1.17–1.43) for DDI AUC ratios and 1.25 (range: 1.02–1.85) for DDI Cmax ratios. This result demonstrated a reasonable optimization of Kcat parameters for BCRP, P-gp and CYP2C9.

#### 3.1.3 Geriatric model

The proposed rosuvastatin PBPK model demonstrated good predictive performance in elderly population, with simulated plasma concentration-time profiles showing good agreement with the observed clinical data (Fig 3, FigS8). Overall model performance was satisfactory, as indicated by a mean relative deviation (MRD) of 1.71.

**Fig 3:**
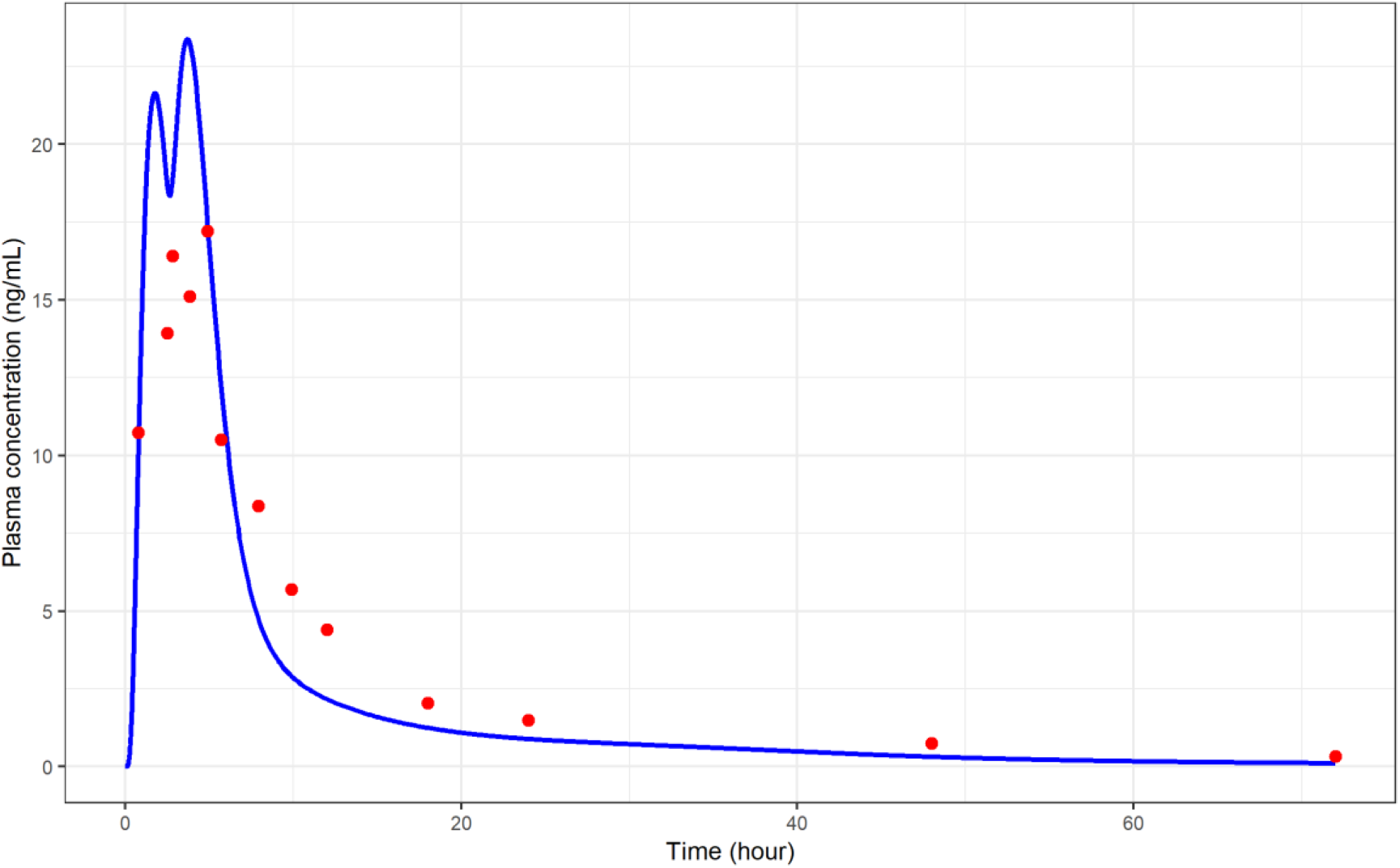
Rosuvastatin plasma concentration–time profiles in geriatric population. Simulations are shown as lines, observed data are shown as dots. Details on administration protocols, study populations, and literature references are listed in table S5.

Comparison of key PK parameters revealed that the predicted AUC was 131 ng·h/mL versus the observed value of 191 ng·h/mL, resulting in a GMFE of 1.46. Similarly, the predicted Cmax was 25 ng/mL compared to the observed Cmax of 18.7 ng/mL, yielding a GMFE of 1.25.

#### 3.1.4 Genetic model

To characterize the impact of ABCG2 genetic variant, the BCRP catalytic turnover rate (Kcat) was optimized to 28 min⁻¹ in subjects with the wildtype genotype of ABCG2 c.421C>A, using clinical PK data. For the wild-type phenotype, the proposed PBPK model demonstrated excellent predictive capability (Fig 4A). Good agreement was observed between simulated and clinically observed plasma concentrations, with strong correlations in the predicted-versus-observed comparisons for AUC, Cmax and plasma concentration (Figs. S9A, S10A, and S11A, respectively). This accuracy was further supported by quantitative metrics, including an overall MRD of 1.35 (1.14– 1.61), and GMFE of 1.23 (1.07–1.36) for AUC and 1.16 (1.03–1.24) for Cmax. Study-specific GMFE and MRD values are summarized in Tables S6 and S7, respectively.

**Fig 4:**
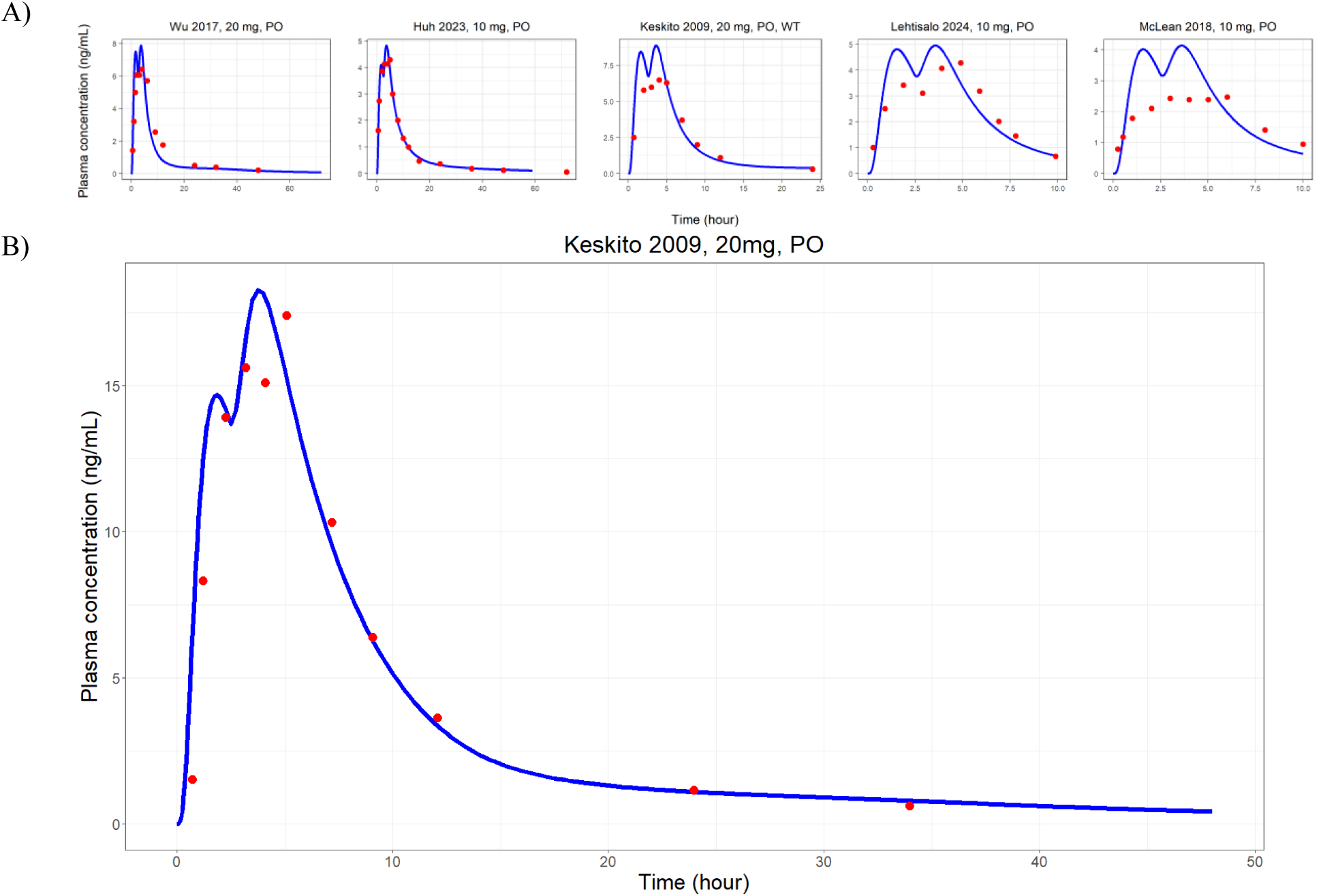
Rosuvastatin plasma concentration–time profiles A) in ABCG2 rs2231142 wildtype genotype and B) in ABCG2 rs2231142 homozygous variant genotype. Simulations are hown as lines, observed data are shown as dots. Details on administration protocols, study populations, and literature references are listed in table S5.

For the ABCG2 variant model, the BCRP Kcat parameter was optimized to 12 min⁻¹ using clinical data. Model adequacy for this genetic subgroup was confirmed through visual predictive checks and predicted-versus-observed plots for AUC and Cmax and plasma concentration (as shown in Figs. S9B, S10B, and S11B) respectively all of which showed close concordance with the clinical data. Quantitatively, the genetic model produced an MRD of 1.2, with GMFE values of 1.10 for AUC and 1.12 for Cmax (Tables S6 and S7), indicating satisfactory predictive performance across all exposure metrics.

In the DDI with rifampin, the model adequately predicted the plasma concentration–time profiles of rosuvastatin both in the presence or absence of rifampin, the predicted concentration was in good agreement with the observed data in subjects with ABCG2 c.421C>A wildtype, as shown in FigS12. In addition, the GMFE index for AUC ratio and Cmax ratio approach 1 as shown in Table S8. This result demonstrated a reasonable value of BCRP Kcat in the genetic model.

### 3.2 Ticagrelor PBPK model

#### 3.2.1 Young adult model

The similar stepwise approach was used for ticagrelor PBPK model development and validation. The final optimized PBPK model parameters for ticagrelor are summarized in Table S9. VPCs are illustrated in Figs5A and 5B, which show good agreement between predicted and observed plasma concentrations in the training and validation datasets, respectively.

The high correlation between predicted and observed AUC, Cmax and plasma concentration across studies in training dataset were depicted in FigsS13A, S14A, S15A, and in validation dataset were illustrated in Figs13B, 14B, 15B. The MRD for PK profiles was 1.56 (range: 1.26–2.08), with 22 of 23 study arms exhibiting an MRD < 2 as shown in table S11. The GMFE values for all AUC and Cmax were within the acceptable two-fold range. Overall, the GMFE for AUC was 1.26 (range: 1.05–1.99) and for Cmax was 1.24 (range: 1.02–1.65), as shown in table S12, indicating robust predictive performance and reliability across multiple validation metrics.

#### 3.2.2 Geriatric model

In the geriatric population, the model also demonstrates good predictive performance as shown by VPC (fig 5C) and strong predicted-versus-observed correlations for AUC, Cmax and plasma concentration in FigS16. Overall, the MRD was 1.53 (range: 1.39-1.68), as shown in table S12 and the GMFE was 1.47 (range: 1.40-1.53) for AUC and 1.22 (range: 1.04-1.40) for Cmax as shown in table S13.

**Fig 5:**
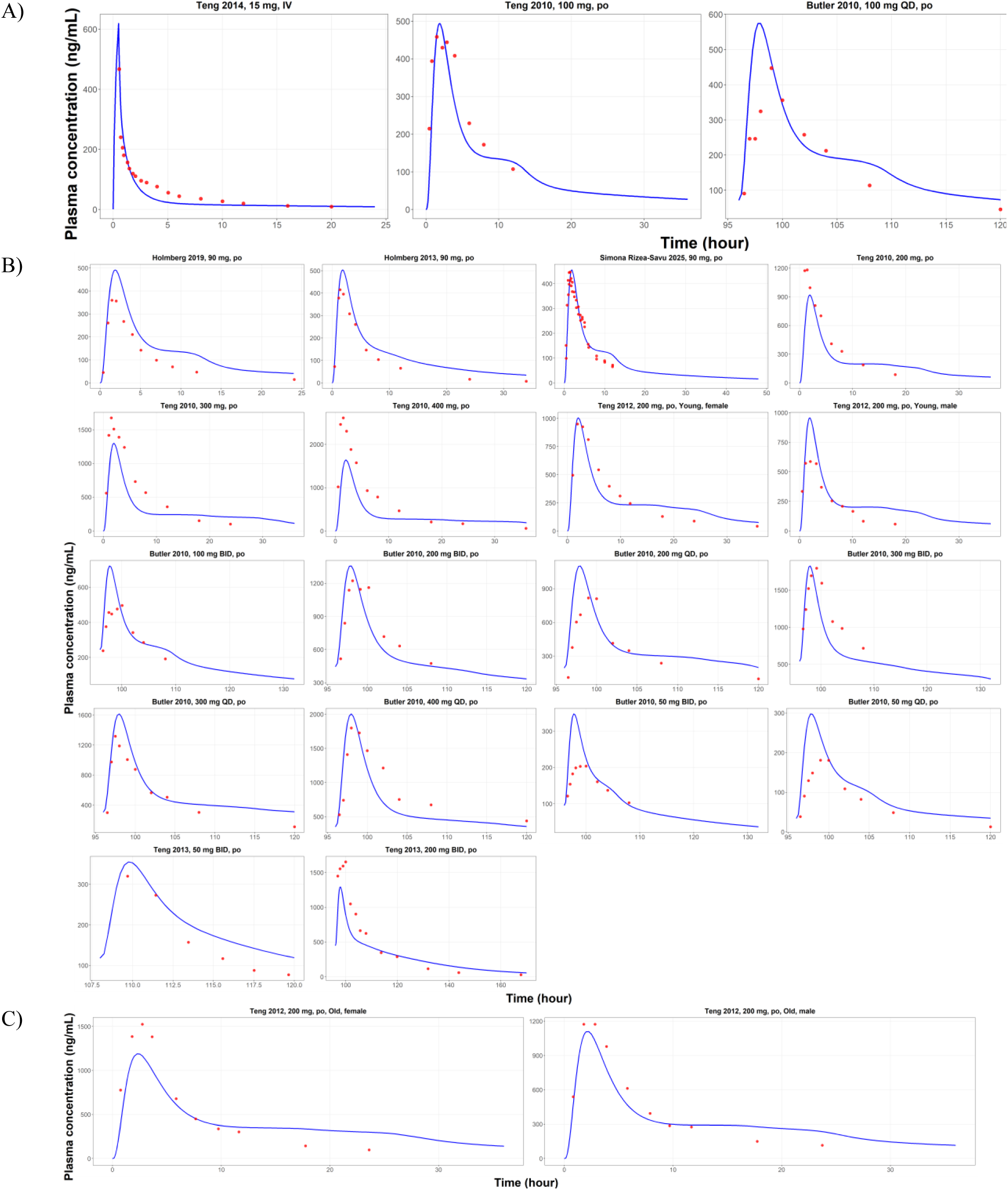
Ticagrelor Plasma Concentration-Time Profiles In A) Training dataset, B) Validation dataset, C) In Elderly Population. Simulated profiles are shown as lines; observed data are shown as dots. Details on administration protocols, study populations, and literature references are listed in table S10.

### 3.3 Ticagrelor-rosuvastatin DDI model in ABCG2 c.421C>A wildtype subjects

With the optimized Ki value of 1.5 × 10^-4^, the model exhibited good predictive performance, as demonstrated by the VPC (Fig 6) and by the direct comparison of predicted and observed DDI ratios for AUC and Cmax. The simulated DDI ratios were in good agreement with the clinical observations, with predicted-versus-observed values of 2.25 versus 2.47 for AUC and 1.92 versus 2.58 for Cmax as shown in Table S14. All predicted DDI ratios fell within both the conventional two-fold error limits and the acceptance thresholds proposed by Guest et al., indicating satisfactory model performance in capturing the magnitude of the observed DDIs.

**Fig 6:**
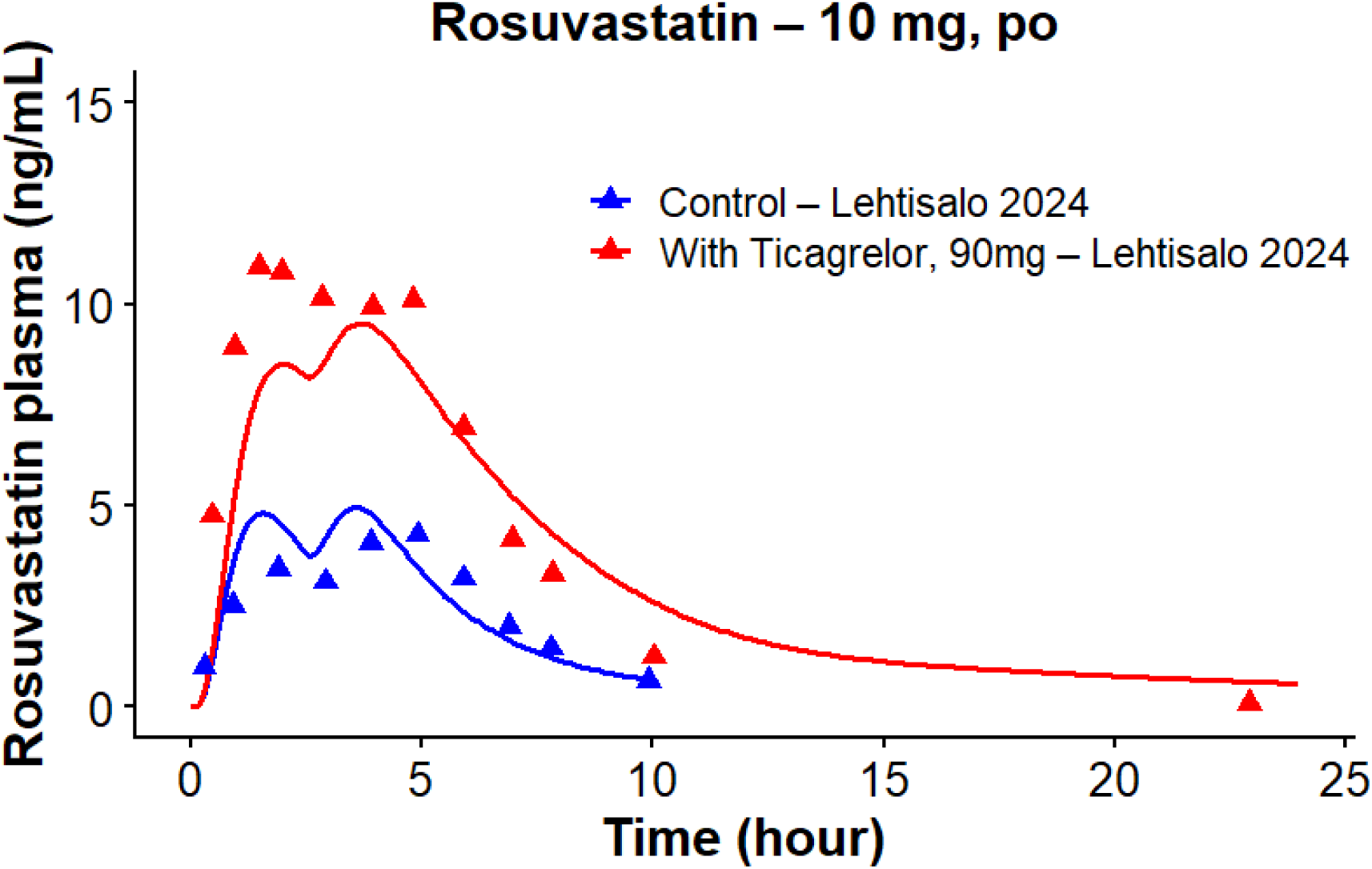
Rosuvastatin Plasma Concentration-Time Profiles Before and During Ticagrelor Coadministration. Simulations are shown as lines, observed data are shown as triangles.

### 3.4 Dose prediction

Young adults: halving the rosuvastatin dose during co-administration with ticagrelor resulted in either similar or moderately altered exposure compared to monotherapy in ABCG2 c.421C>A wild-type and homozygous genotypes, respectively, supporting a half-dose initiation strategy in this population, as shown in table 1. However, close monitoring is warranted in ABCG2 c.421C>A wild-type individuals, as the mean fold change (1.44) slightly exceeds the predefined upper threshold (1.25).

**Table 1:** Dose prediction of rosuvastatin in adult population with ABCG2 c.421C>A wildtype genotype.

| Monotherapy<br>Dose | Cmax<br>(Monotherapy) | Cmax with<br>Ticagrelor<br>(Half Dose) | Cmax<br>Fold<br>Change | AUClast<br>(Monotherapy) | AUClast<br>with<br>Ticagrelor<br>(Half Dose) | AUClast<br>Fold<br>Change |
| --- | --- | --- | --- | --- | --- | --- |
| 5 mg | 1.97 | 2.35 | 1.19 | 13.88 | 18.99 | 1.37 |
| 10 mg | 3.65 | 4.49 | 1.23 | 25.85 | 37.57 | 1.45 |
| 20 mg | 7.69 | 8.76 | 1.14 | 58.68 | 80.49 | 1.37 |
| 40 mg | 16.87 | 17.87 | 1.06 | 119.94 | 188.24 | 1.57 |
|  |  | Mean fold<br>change | 1.16 |  | Mean fold<br>change | 1.44 |

**Table 2:**
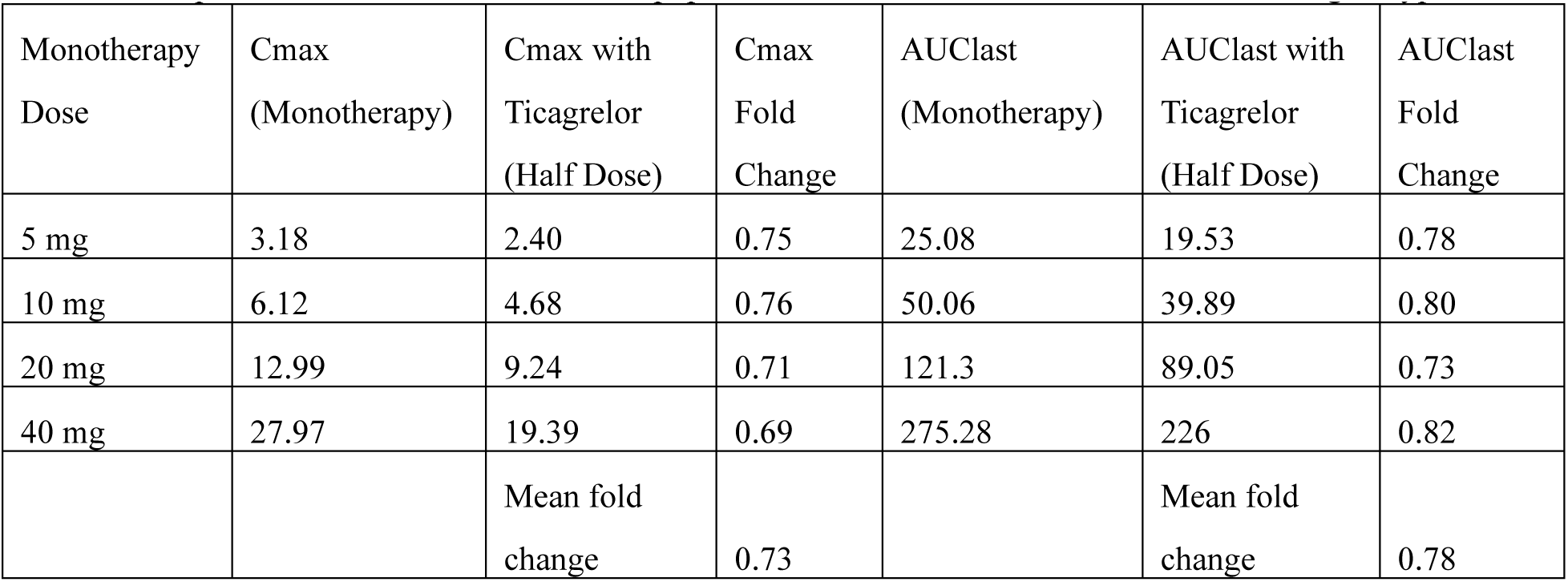
Dose prediction of rosuvastatin in adult population with BCRP ABCG2 c.421C>A variant genotype.

Geriatric population: dose recommendations varied by genotype. For wild-type individuals, halving the dose still produced a significant AUC increase compared to monotherapy, necessitating a four-fold lower starting dose as shown in table 3. For variant carriers, a half-dose reduction achieved comparable exposure to monotherapy, supporting a half-dose initiation strategy in this subgroup as well as shown in table 4.

**Table 3:** Dose prediction of rosuvastatin in elderly population with ABCG2 c.421C>A wildtype genotype.

| Monotherapy<br>Dose | Cmax<br>(Monotherapy) | Cmax with Ticagrelor |  | Cmax Fold Change |  | AUClast<br>(Monotherapy) | AUClast with<br>Ticagrelor |  | AUClast Fold Change |  |
| --- | --- | --- | --- | --- | --- | --- | --- | --- | --- | --- |
|  |  | Half Dose | One<br>fourth<br>dose | Half Dose | One<br>fourth<br>dose |  | Half Dose | One<br>fourth<br>dose | Half Dose | One<br>fourth<br>dose |
| 5 mg | 2.71 | 3.92 | 1.95 | 1.45 | 0.72 | 14.35 | 32.03 | 14.93 | 2.23 | 1.04 |
| 10 mg | 5.00 | 7.35 | 3.92 | 1.47 | 0.78 | 26.86 | 61.40 | 32.03 | 2.29 | 1.19 |
| 20 mg | 10.07 | 13.61 | 7.35 | 1.35 | 0.73 | 55.29 | 117.85 | 61.40 | 2.13 | 1.11 |
| 40 mg | 21.74 | 25.97 | 13.61 | 1.19 | 0.63 | 129.03 | 237.82 | 117.85 | 1.84 | 0.91 |
|  |  |  | Mean fold<br>change | 1.37 | 0.71 |  |  | Mean fold<br>change | 2.12 | 1.06 |

**Table 4:** Dose prediction of rosuvastatin in elderly population with BCRP ABCG2 c.421C>A variant genotype.

| Monotherapy<br>Dose | Cmax<br>(Monotherapy) | Cmax with<br>Ticagrelor<br>(Half Dose) | Cmax<br>Fold<br>Change | AUClast<br>(Monotherapy) | AUClast<br>with<br>Ticagrelor<br>(Half Dose) | AUClast Fold<br>Change |
| --- | --- | --- | --- | --- | --- | --- |
| 5 mg | 4.31 | 4.1 | 0.95 | 29.58 | 35 | 1.18 |
| 10 mg | 8.18 | 7.73 | 0.94 | 56.69 | 68.59 | 1.21 |
| 20 mg | 16.58 | 14.39 | 0.87 | 119.83 | 134.21 | 1.12 |
| 40 mg | 36.8 | 27.76 | 0.75 | 286.4 | 278 | 0.97 |
|  |  | Mean fold<br>change | 0.88 |  | Mean fold<br>change | 1.12 |

## 4. Discussion

In this study, optimal rosuvastatin dose during co-administration with ticagrelor in terms of both age and ABCG2 c.421C>A genotype was predicted using PBPK modelling approach. In young adults, a half-dose reduction appears appropriate regardless of ABCG2 genotypes. In the elderly, a four-fold dose reduction is required for wild-type individuals, whereas variant carriers can safely initiate with a half-dose reduction. These findings provide genotype- and age-specific dose regimen to minimize toxicity risk without compromising efficacy.

Our simulations showed that ticagrelor markedly increased rosuvastatin exposure in individuals carrying the wild-type ABCG2 c.421C allele, whereas only a modest increase in PK exposure in individuals’ carriers of the ABCG2 c.421A variant, which is associated with reduced transporter function. This observation is mechanistically plausible. In individuals with the wild-type ABCG2 genotype, BCRP transporter activity is highest, indicating this efflux transporter as a key determinant of rosuvastatin PK. Consequently, inhibition of BCRP by ticagrelor leads to a substantial increase in systemic exposure. Conversely, in carriers of the ABCG2 c.421C>A homozygous variant, baseline BCRP activity is reduced, rendering rosuvastatin PK less dependent on this pathway. As a result, BCRP inhibition produces a comparatively smaller relative change in exposure in this genotype group.

This genotype driven DDI pattern somehow mirrors clinical observations reported for CYP2C19-mediated interactions, such as the lansoprazole–fluvoxamine interaction, where co-administration of the strong CYP2C19 inhibitor fluvoxamine increased lansoprazole exposure by nearly four-fold in CYP2C19 extensive metabolizers, while producing minimal additional exposure increase in CYP2C19 poor metabolizers[25]. In both cases, pharmacological inhibition exerts its largest effect in individuals with preserved baseline pathway activity, whereas genetic loss-of-function variants attenuate the apparent magnitude of DDI due to pre-existing functional impairment of the affected enzyme or transporter. These findings support the concept that DDI are often the dominant determinant of exposure changes, while genetic polymorphisms primarily modulate baseline PK and susceptibility to interaction-induced phenoconversion.

In addition to genetic factors, old age was reported as possible multifactorial causes for rhabdomyolysis following concomitant use of rosuvastatin and ticagrelor[26]. In addition, ticagrelor exposure was reported to have been around 1.5 folds higher in elderly versus the young[14]. Therefore, a dose optimization analysis remains to answer in this understudied population. In the geriatric population, an age-related 40% reduction in CYP2C9 activity was incorporated into the model. This further diminishes an alternative clearance pathway, thereby amplifying the relative contribution of BCRP to overall rosuvastatin disposition. The combined effect of reduced metabolic clearance and high transporter activity explains why, even after halving the dose, rosuvastatin exposure remained significantly elevated in elderly adults with the ABCG2 wildtype genotype compared to monotherapy. These findings support the need for a more conservative, four-fold dose reduction when initiating rosuvastatin concomitantly with ticagrelor in this vulnerable subpopulation.

The optimized Ki is far lower than the reported in-vitro vesicular value. However, this discrepancy most likely reflects fundamental differences between isolated transporter systems and the physiological enterocyte environment. Vesicular assays measure inhibition under artificial membrane conditions with transporter overexpression and nominal incubation concentrations, whereas the PBPK model accounts for intracellular unbound concentrations within polarized enterocytes.

Notably, the rosuvastatin PBPK model demonstrated preserved predictive performance in an independent rifampin interaction scenario. This robustness suggests that the parameter represents an apparent in-vivo inhibition constant incorporating system-level scaling factors rather than an artifact of model overfitting. We further hypothesized that the highly protein-bound nature of ticagrelor may contribute to this difference, as the model predicted a low maximum unbound intracellular concentration in small-intestinal enterocytes (approximately 2 nmol/L). Under such conditions, small differences in effective intracellular exposure may translate into substantial shifts in the apparent inhibition constant required to recover the observed clinical AUC ratio.

The predicted PK profiles were also consistent with findings reported in other clinical studies. The half-life of rosuvastatin has been reported to be approximately 30% higher in elderly subjects, which is consistent with an approximate 50% reduction in oral clearance compared with young adults[27, 28]. In addition, plasma concentrations of rosuvastatin have been reported to be approximately two-fold higher in subjects receiving ticagrelor compared with those receiving rosuvastatin alone or in combination with other drugs[1, 29]. Similarly, the AUC and Cmax of rosuvastatin have been shown to increase up to two-fold in individuals with the ABCG2 c.421C>A homozygous variant compared with heterozygous or wild-type genotypes[7, 30].

Our study has a few key limitations. First, due to the limited available clinical data, the PBPK model for scenarios involving elderly populations, individuals carrying the ABCG2 c.421C>A variant, and the co-administration of ticagrelor with rosuvastatin was evaluated using only a single clinical study. In these scenarios, the model was mechanistically optimized based on clinical data. Due to insufficient demographic and clinical dose information, as well as potential differences in ethnicity across studies, these datasets could not be used for model validation. Second, the model was developed and evaluated primarily using data from Caucasian populations. Given the known ethnic differences in rosuvastatin PK, further studies are warranted to investigate the influence of ticagrelor on rosuvastatin exposure in other populations, such as Asian and African populations.

## Declarations

### Author Contributions

H.D.N., J.D., J.T., designed the research. H.D.N., J.D., performed the research. H.D.N., J.D., analyzed the data. H.D.N., V.H.P., J.D., wrote the manuscript. J.T., reviewed the manuscript.

### Data Availability Statement

The datasets generated during and/or analyzed during the current study are available from the corresponding author on reasonable request.

### Ethical Approval

Not applicable

### Consent to participate

Not applicable

### Consent for publication

Not applicable.

## ACKNOWLEDGEMENTS

This study was sponsored by Wellcome Trust (Grant No.). The funding source provided financial support but had no role in the study design, data collection, analysis or interpretation, nor in the writing of the manuscript or the decision to submit it for publication.

