## Supplementary for "PBPK Model-Based Prediction of Optimal Rosuvastatin Dosing with Concomitant Ticagrelor in Elderly Patients Considering ABCG2 Genetic Variant"

**Table legend**

Table S1: The input parameters for the development of rosuvastatin PBPK model.

Table S2: Mean relative deviation (MRD) of rosuvastatin plasma concentration predictions in base model

Table S3: Predicted and observed rosuvastatin AUClast and Cmax values with geometric mean fold errors (GMFEs) in base model.

Table S4: Predicted and Observed Rifampicin-Rosuvastatin DDI Ratios for AUC_last_ and C_max_ with Geometric Mean Fold Errors (GMFEs).

Table S5: Clinical studies used for rosuvastatin ABCG2 c.421C>A genetic PBPK model development and validation in geriatric population and subjects with wildtype and homozygous genotype of ABCG2 c.421C>A.

Table S6: Mean relative deviation (MRD) of rosuvastatin plasma concentration predictions in ABCG2 genetic model.

Table S7: Predicted and observed rosuvastatin AUClast and Cmax values with geometric mean fold errors (GMFEs) in *ABCG2* c.421C>A wildtype and homozygous variant genotype.

Table S8: Predicted and Observed Rifampicin-Rosuvastatin DDI Ratios for AUC_last_ and C_max_ with Geometric Mean Fold Errors (GMFEs) in subjects with wild type genotype of *ABCG2* c.421C>A.

Table S9: The input parameters for the development of ticagrelor PBPK model.

Table S10: Clinical studies used for ticagrelor PBPK model development and validation.

Table S11: Mean relative deviation (MRD) of ticagrelor plasma concentration prediction.

Table S12: Predicted and observed ticagrelor AUClast and Cmax values with geometric mean fold errors (GMFEs) in base model.

Table S13: Mean relative deviation (MRD) of ticagrelor plasma concentration prediction in geriatric population.

Table S14: Predicted and observed rosuvastatin AUClast and Cmax values with geometric mean fold errors (GMFEs) in geriatric population.

Table S15: Predicted and Observed Ticagrelor-Rosuvastatin DDI Ratios for AUC_last_ and C_max_ with Geometric Mean Fold Errors (GMFEs) in subjects with wild type genotype of *ABCG2* c.421C>A.

**Figure legends**

FigS1: Predicted versus observed fraction of rosuvastatin excreted in urine. Simulations are shown as lines, observed data are shown as dots. Details on administration protocols, study populations, and literature references are listed in previous study(1).

FigS2: Predicted versus observed rosuvastatin AUClast values for the training (a) and test (b) datasets in the base model. Solid lines represent the line of identity. Dotted lines indicate 1.25-fold deviation; dashed lines indicate 2-fold deviation. Details on administration protocols, study populations, and literature references are listed in previous study(1).

FigS3: Predicted versus observed rosuvastatin Cmax values for the training (a) and test (b) datasets in the base model. Solid lines represent the line of identity. Dotted lines indicate 1.25-fold deviation; dashed lines indicate 2-fold deviation. Details on administration protocols, study populations, and literature references are listed in previous study(1).

FigS4: Predicted versus observed rosuvastatin plasma concentration values for the training (a) and test (b) datasets in the base model. Solid lines represent the line of identity. Dotted lines indicate 1.25-fold deviation; dashed lines indicate 2-fold deviation. Details on administration protocols, study populations, and literature references are listed in previous study(1).

FigS5: Rosuvastatin plasma concentration-time profiles before and during rifampicin coadministration.Simulations are shown as lines, observed data are shown as dots. Details on administration protocols, study populations, and literature references are listed in previous study(1).

FigS6: Rosuvastatin fraction excreted in urine profiles, before and during oral rifampicin co-administration. Simulations are shown as lines, observed data are shown as dots. Details on administration protocols, study populations, and literature references are listed in previous study(1).

FigS7: Predicted versus observed rifampicin-rosuvastatin DDI AUClast ratios and DDI Cmax ratios in base model. The straight solid lines ( ) mark the line of identity. Dotted lines ( ) indicate 1.25-fold, dashed lines ( ) indicate 2-fold deviation. The curved solid lines illustrate the DDI prediction success limits suggested by Guest et al(2).

FigS8: Predicted versus observed rosuvastatin plasma concentration in model for geriatric population. Solid lines represent the line of identity. Dotted lines indicate 1.25-fold deviation; dashed lines indicate 2-fold deviation. Details on administration protocols, study populations, and literature references are listed in table S5.

FigS9: Predicted versus observed rosuvastatin AUClast values for the *ABCG2* c.421C>A wild-type (a) and variant (b) datasets in genetic model. Solid lines represent the line of identity. Dotted lines indicate 1.25-fold deviation; dashed lines indicate 2-fold deviation. Details on administration protocols, study populations, and literature references are listed in table S5.

FigS10: Predicted versus observed rosuvastatin Cmax values for the *ABCG2* c.421C>A wild-type (a) and variant (b) datasets in genetic model. Solid lines represent the line of identity. Dotted lines indicate 1.25-fold deviation; dashed lines indicate 2-fold deviation. Details on administration protocols, study populations, and literature references are listed in table S5.

FigS11: Predicted versus observed rosuvastatin plasma concentration values for the *ABCG2* c.421C>A wild-type (a) and variant (b) datasets in genetic model. Solid lines represent the line of identity. Dotted lines indicate 1.25-fold deviation; dashed lines indicate 2-fold deviation. Details on administration protocols, study populations, and literature references are listed in table S5.

FigS12: Rosuvastatin plasma concentration-time profiles, before and during rifampicin coadministration in subjects with *ABCG2* c.421C>A wild-type genotype. Simulations are shown as lines, observed data are shown as dots.

FigS13: Predicted versus observed ticagrelor AUClast values for the training (a) and test (b) datasets in the base model. Solid lines represent the line of identity. Dotted lines indicate 1.25-fold deviation; dashed lines indicate 2-fold deviation. Details on administration protocols, study populations, and literature references are listed in table S10.

FigS14: Predicted versus observed ticagrelor Cmax values for the training (a) and test (b) datasets in the base model. Solid lines represent the line of identity. Dotted lines indicate 1.25-fold deviation; dashed lines indicate 2-fold deviation. Details on administration protocols, study populations, and literature references are listed in table S10.

FigS15: Predicted versus observed ticagrelor plasma concentration values for the training (a) and test (b) datasets in the base model. Solid lines represent the line of identity. Dotted lines indicate 1.25-fold deviation; dashed lines indicate 2-fold deviation. Details on administration protocols, study populations, and literature references are listed in table S10.

FigS16: Predicted versus observed ticagrelor AUClast(a), Cmax (b) and plasma concentration (c) values for the geriatric population. Solid lines represent the line of identity. Dotted lines indicate 1.25-fold deviation; dashed lines indicate 2-fold deviation. Details on administration protocols, study populations, and literature references are listed in table S9.

Table S1: The input parameters for the development of rosuvastatin PBPK model.

| Parameter | Value in the previous model | Value in the recent model | Unit |
| --- | --- | --- | --- |
| Molecular weight (MW) | 481.54 | No change | g/mol |
| pKa (acid) | 4.3 | No change | – |
| Solubility (water) | 7.8 | No change | g/L |
| logP | −0.33 | No change | – |
| Fraction unbound (fu) | 11.5 | No change | % |
| BCRP Km | 2.02 | 17 | µmol/L |
| BCRP kcat | 1.01 | 19 | 1/min |
| CYP2C9 CLspec | 1.04E-03 | 1.84E-03 | L/µmol·min |
| OAT3 Km | 7.40 | No change | µmol/L |
| OAT3 kcat | 66851.33 | No change | 1/min |
| OATP1B1/1B3 Km | 0.80 | No change | µmol/L |
| OATP1B1/1B3 kcat | 1124.12 | No change | 1/min |
| OATP2B1 Km | 6.42 | No change | µmol/L |
| OATP2B1 kcat | 5.00 | No change | 1/min |
| P-gp Km | 203.00 | No change | µmol/L |
| P-gp kcat | 36.07 | 18 | 1/min |
| GFR fraction | 1.00 | No change | – |
| Enterohepatic circulation (EHC) continuous fraction | 1.00 | No change | – |
| Partition coefficients | Diverse | No change | – |
| Cellular permeability | 1.34E-06 | 3.24E-06 | cm/min |
| Intestinal permeability | 1.51E-06 | No change | cm/min |
| P(intracellular → interstitial), small intestine | 4.31E-04 | No change | cm/min |
| P(intracellular → interstitial), large intestine | 3.66E-06 | No change |  |
| Tablet dissolution Weibull shape | 1.77 | No change |  |
| Tablet dissolution Weibull time | 6.25 | No change | min |

Table S2: Mean relative deviation (MRD) of rosuvastatin plasma concentration predictions in base model

| Route | Dose (mg) | MRD | Reference |
| --- | --- | --- | --- |
| iv, bolus | 0.00174 | 1.83 | Billington et al. 2019 |
| iv, 4 h | 8 | 1.63 | Martin et al. 2003c |
| Oral, sol | 0.05 | 1.40 | Prueksaritanont et al. 2017 |
| Oral, - | 5 | 2.24 | Prueksaritanont et al. 2014 |
| Oral, caps | 10 | 1.48 | Cooper et al. 2003b |
| Oral, - | 10 | 1.84 | Coss et al. 2016 |
| Oral, tab | 10 | 1.56 | Csonka et al. 2019 |
| Oral, tab | 10 | 1.41 | Huguet et al. 2016 |
| Oral, tab | 10 | 1.43 | Martin et al. 2003b |
| Oral, tab | 10 | 1.46 | Stopfer et al. 2016 |
| Oral, tab | 10 | 1.45 | Stopfer et al. 2018a |
| Oral, tab | 10 | 1.46 | Stopfer et al. 2018b |
| Oral, tab | 10 | 1.71 | Wiebe et al. 2020 |
| Oral, tab | 10 | 1.69 | Wiebe et al. 2020 |
| Oral, - | 10 QD | 1.32 | Kosoglou et al. 2004 |
| Oral, - | 10 QD | 1.31 | Martin et al. 2002b |
| Oral, caps, qd | 20 QD | 1.38 | FDA 2002 |
| Oral, - | 20 QD | 1.24 | FDA 2018 |
| Oral, - | 20 QD | 1.35 | Lee et al. 2018 |
| Oral, - | 20 | 1.61 | Birmingham et al. 2015 |
| Oral, tab | 20 | 2.18 | Edwards et al. 2017 |
| Oral, caps | 20 | 1.32 | FDA 2002 |
| Oral, - | 20 | 1.55 | Jones et al. 2020 |
| Oral, - | 20 | 1.32 | Lee et al. 2018 |
| Oral, tab | 20 | 2.24 | Martin et al. 2003b |
| Oral, sol | 20 | 1.51 | Martin et al. 2003d |
| Oral, - | 20 | 1.39 | Martin et al. 2016 |
| Oral, - | 20 | 1.30 | Willis et al. 2020 |
| Oral, - | 40 QD | 1.31 | Gosai et al. 2008 |
| Oral, tab | 40 | 1.27 | Gidal et al. 2017 |
| Oral, - | 40 | 1.42 | Lee et al. 2005 |
| Oral, caps | 40, female | 1.51 | Martin et al. 2002a |
| Oral, caps | 40, male | 1.36 | Martin et al. 2002a |
| Oral, tab | 40 | 28 | Martin et al. 2003b |
| Oral, tab | 40 | 1.54 | Martin et al. 2003c |
| Oral, tab | 80 | 1.72 | Cooper et al. 2002 |
| Oral, tab | 80 | 1.95 | Cooper et al. 2003a |
| Oral, tab | 80 | 1.74 | Cooper et al. 2003b |
| Oral, tab | 80 | 1.93 | Cooper et al. 2003c |
| Oral, tab | 80 | 2.23 | Martin et al. 2003b |
| Oral, tab | 80 | 1.43 | Schneck et al. 2004 |

Table S3 Predicted and observed rosuvastatin AUClast and Cmax values with geometric mean fold errors (GMFEs) in base model.

| Route | Formulation | Dose  (mg) | Predicted AUC (ng*h/ml) | Observed AUC  (ng*h/ml) | GMFE AUC | Predicted Cmax  (ng/ml) | Observed Cmax  (ng/ml) | GMFE Cmax | Reference |
| --- | --- | --- | --- | --- | --- | --- | --- | --- | --- |
| Intravenous bolus | - | 0.00174 | 0.0216 | 0.03 | 1.38 | – |  |  | Billington et al. 2019 |
| Intravenous 4 h infusion | - | 8 | 179.25 | 163.14 | 1.09 | – |  |  | Martin et al. 2003c |
| Oral | solution | 0.05 b | 0.2264 | 0.21 | 1.07 | 0.0346 | 0.03 | 1.15 | Prueksaritanont et al. 2017 |
| Oral | – | 5 | 16.66 | 37.46 | 2.24 | 2.34 | 4.59 | 1.95 | Prueksaritanont et al. 2014 |
| Oral | capsule | 10 | 31.92 | 44.07 | 1.38 | 4.29 | 5.37 | 1.25 | Cooper et al. 2003b |
| Oral | tablet | 10 | 36.87 | 55.45 | 1.50 | 4.245 | 5.13 | 1.20 | Csonka et al. 2019 |
| Oral | tablet | 10 | 34.83 | 33.05 | 1.05 | 4.642 | 3.58 | 1.29 | Huguet et al. 2016 |
| Oral | tablet | 10 | 31.58 | 32.92 | 1.04 | 4.243 | 3.73 | 1.13 | Martin et al. 2003b |
| Oral | tablet | 10 | 31.75 | 41 | 1.29 | 4.101 | 4.48 | 1.09 | Stopfer et al. 2016 |
| Oral | tablet | 10 | 33.18 | 46.26 | 1.393 | 4.05 | 4.97 | 1.22 | Stopfer et al. 2018a |
| Oral | tablet | 10 | 34.18 | 42.22 | 1.23 | 4.188 | 4.11 | 1.01 | Stopfer et al. 2018b |
| Oral | tablet | 10 | 33.43 | 54.88 | 1.64 | 4.095 | 5.12 | 1.25 | Wiebe et al. 2020 |
| Oral | tablet | 10 | 34.46 | 54.88 | 1.59 | 4.24 | 5.12 | 1.20 | Wiebe et al. 2020 |
| Oral | – | 10 | 39.6 | 59.54 | 1.50 | 4.62 | 5.07 | 1.09 | Coss et al. 2016 |
| Oral | solution | 20 | 68.5 | 63.22 | 1.08 | 8.27 | 6.07 | 1.36 | Martin et al. 2003d |
| Oral | capsule | 20 | 73.04 | 89.29 | 1.22 | 8.94 | 10.31 | 1.15 | FDA et al. 2003 |
| Oral | tablet | 20 | 82.92 | 123.77 | 1.49 | 9.52 | 11.01 | 1.15 | Edwards et al. 2017 |
| Oral | tablet | 20 | 63.4 | 54.22 | 1.16 | 8.55 | 6.96 | 1.22 | Martin et al. 2003b |
| Oral | tablet | 20 | 59.38 | 71.63 | 1.20 | 7.87 | 6.43 | 1.22 | Wu et al. 2017 |
| Oral | – | 20 | 93.16 | 93.77 | 1.00 | 10.93 | 8.23 | 1.32 | Birmingham et al. 2015 |
| Oral | – | 20 | 72.67 | 98.51 | 1.35 | 10.23 | 10.1 | 1.01 | Jones et al. 2020 |
| Oral | – | 20 | 62.9 | 84.63 | 1.34 | 8.86 | 8.83 | 1.00 | Lee et al. 2018 |
| Oral | – | 20 | 70.87 | 101.02 | 1.42 | 8.32 | 10.26 | 1.23 | Martin et al. 2016 |
| Oral | – | 20 | 55.67 | 65.9 | 1.18 | 9.08 | 8.88 | 1.02 | Willis et al. 2020 |
| Oral | capsule | 40 | 220.3 | 141.49 | 1.55 | 30.96 | 16.99 | 1.82 | Martin et al. 2002a |
| Oral | capsule | 40 | 122.76 | 158.63 | 1.29 | 19.66 | 20.5 | 1.04 | Martin et al. 2002a |
| Oral | tablet | 40 | 184.52 | 190.05 | 1.02 | 20.45 | 18.03 | 1.13 | Gidal et al. 2017 |
| Oral | tablet | 40 | 161.36 | 93.84 | 1.71 | 18.73 | 9.5 | 1.97 | Martin et al. 2003b |
| Oral | tablet | 40 | 169.06 | 158.94 | 1.06 | 19.07 | 18.07 | 1.05 | Martin et al. 2003c |
| Oral | – | 40 | 168.41 | 234.87 | 1.39 | 18.99 | 23.35 | 1.22 | Lee et al. 2005 |
| Oral | tablet | 80 | 386.64 | 318.12 | 1.21 | 43.18 | 37.83 | 1.14 | Cooper et al. 2002 |
| Oral | tablet | 80 | 459.45 | 311.14 | 1.47 | 50.50 | 36.2 | 1.39 | Cooper et al. 2003a |
| Oral | tablet | 80 | 399.73 | 392.78 | 1.01 | 44.21 | 53.37 | 1.20 | Cooper et al. 2003b |
| Oral | tablet | 80 | 395.97 | 241.93 | 1.63 | 44.09 | 30.88 | 1.42 | Cooper et al. 2003c |
| Oral | tablet | 80 | 393.07 | 257.68 | 1.52 | 43.80 | 29.29 | 1.49 | Martin et al. 2003b |
| Oral | tablet | 80 | 395.23 | 404.47 | 1.02 | 50.08 | 45.19 | 1.10 | Schneck et al. 2004 |
| Oral | – (qd) | 10 | 40.5 | 46.58 | 1.15 | 5.19 | 4.43 | 1.17 | Kosoglou et al. 2004 |
| Oral | – (qd) | 10 | 43.8 | 49.73 | 1.13 | 4.85 | 4.42 | 1.09 | Martin et al. 2002b |
| Oral | capsule (qd) | 20 | 107.2 | 100.98 | 1.06 | 10.26 | 9.52 | 1.07 | FDA et al. 2003 |
| Oral | – (qd) | 20 | 129 | 114.91 | 1.12 | 15.65 | 11.03 | 1.41 | FDA et al. 2018 |
| Oral | – (qd) | 20 | 86.5 | 123.9 | 1.43 | 10.18 | 11.77 | 1.15 | Lee et al. 2018 |
| Oral | – (qd) | 40 | 188.2 | 219.39 | 1.16 | 22.79 | 24.1 | 1.05 | Gosai et al. 2008 |
|  |  |  |  | Mean GMFE AUC | 1.30 |  | Mean GMFE Cmax | 1.24 |  |

Table S4: Predicted and Observed Rifampicin-Rosuvastatin DDI Ratios for AUC_last_ and C_max_ with Geometric Mean Fold Errors (GMFEs).

| Rifampin | Rosuvastatin | Dose gap [h] | n | DDI AUC _last_ ratio | | | DDI C_max_ ratio | | | Reference |
| --- | --- | --- | --- | --- | --- | --- | --- | --- | --- | --- |
|  |  |  |  | Pred | Obs | GMFE | Pred | Obs | GMFE |  |
| 600 mg, iv, 0.5 h | 5.0 mg, oral, - | 0.5 | 8 | 4.1 | 4.83 | 1.17 | 9.06 | 8.88 | 1.02 | Prucksaritanont et al. 2014 |
| 600 mg, oral, caps | 0.05 mg a, oral, sol | 0 | 12/8 | 4.1 | 5.88 | 1.43 | 11.6 | 12.41 | 1.06 | Prucksaritanont et al. 2017 |
| 600 mg, oral, - | 5.0 mg, oral, - | 0 | 8 | 4.92 | 3.69 | 1.33 | 11 | 5.94 | 1.85 | Prucksaritanont et al. 2014 |
| 600 mg, oral, tab | 10.0 mg b, oral, tab | 0 | 11 | 4.3 | 3.52 | 1.22 | 10.6 | 9.96 | 1.06 | Wiebe et al. 2020 |

Table S5: Clinical studies used for rosuvastatin ABCG2 c.421C>A genetic PBPK model development and validation in geriatric population and subjects with wildtype and homozygous genotype of ABCG2 c.421C>A.

| Author year | Models | ABCG2 c.421C>A genotype | N | Age (year) | Weight (kg) | Gender (% female) | Dose  (mg) | Route |
| --- | --- | --- | --- | --- | --- | --- | --- | --- |
| Martin et al., 2012 | Elderly | Not reported | 16 | 67.7 | 71.8 | 50 | 40 | Oral |
| Huh et al., 2023(3) | Genetic | Wildtype | 12 | 38.7 | 74.9 | 16.7 | 10 | Oral |
| Keskito et al., 2009(3) | Genetic | Wildtype | 16 | 22 | 67 | 50 | 20 | Oral |
| Lehtisalo et al., 2024(4) | Genetic | Wildtype | 9 | 24 | 64 | 66 | 10 | Oral |
| McLean et al., 2018(5) | Genetic | Wildtype | 11 | 22 | 79 | 45 | 10 | Oral |
| Wu et al., 2017(6) | Genetic | Wildtype | 7 | 43 | 68 | 43 | 20 | Oral |
| Keskito et al., 2009(3) | Genetic | Homozygous variant | 4 | 21 | 58 | 75 | 20 | Oral |

Table S6: Mean relative deviation (MRD) of rosuvastatin plasma concentration predictions in ABCG2 genetic model.

| Route | ABCG2 c.421C>A genotype | Dose (mg) | MRD | Reference |
| --- | --- | --- | --- | --- |
| Oral | Wildtype | 10 | 1.14 | Huh et al., 2023 |
| Oral | Wildtype | 20 | 1.27 | Keskito et al., 2009 |
| Oral | Wildtype | 10 | 1.36 | Lehtisalo et al., 2024 |
| Oral | Wildtype | 10 | 1.61 | McLean et al., 2018 |
| Oral | Wildtype | 20 | 1.38 | Wu et al., 2017 |
| Oral | Homozygous variant | 20 | 1.28 | Keskito et al., 2009 |

Table S7: Predicted and observed rosuvastatin AUClast and Cmax values with geometric mean fold errors (GMFEs) in *ABCG2* c.421C>A wildtype and homozygous variant genotype.

| Route | *ABCG2* c.421C>A | Formulation | Dose  (mg) | Predicted AUC  (ng*h/ml) | Observed AUC  (ng*h/ml) | GMFE AUC | Predicted Cmax | Observed Cmax | GMFE Cmax | ID |
| --- | --- | --- | --- | --- | --- | --- | --- | --- | --- | --- |
| Oral | Wildtype | tablet | 10 | 49.35 | 43.86 | 1.13 | 4.83 | 4.49 | 1.08 | Huh et al. 2023 |
| Oral | Wildtype | tablet | 20 | 84.3 | 62.3 | 1.35 | 8.88 | 7.06 | 1.26 | Keskitalo et al. 2009 |
| Oral | Wildtype | tablet | 10 | 26.88 | 28.9 | 0.93 | 4.7 | 5.30 | 0.89 | Lehtisalo et al. 2023 |
| Oral | Wildtype | tablet | 10 | 22.93 | 18.3 | 1.25 | 3.9 | 2.90 | 1.34 | McLean et al. 2018 |
| Oral | Wildtype | tablet | 20 | 56.76 | 77.2 | 0.74 | 7.88 | 7.60 | 1.04 | Wu et al. 2017 |
| Oral | Homozygous variant | tablet | 20 | 168.845 | 152.2 | 1.11 | 18.30 | 16.33 | 1.12 | Keskitalo et al. 2009 |

Table S8: Predicted and Observed Rifampicin-Rosuvastatin DDI Ratios for AUC_last_ and C_max_ with Geometric Mean Fold Errors (GMFEs) in subjects with wild type genotype of *ABCG2* c.421C>A.

| Rifampin | Rosvastatin | Dose gap [h] | n | DDI AUC _last_ ratio | | | DDI C_max_ ratio | | | Reference |
| --- | --- | --- | --- | --- | --- | --- | --- | --- | --- | --- |
|  |  |  |  | Pred | Obs | GMFE | Pred | Obs | GMFE |  |
| 600 mg, iv, 0.5 h | 20.0 mg, oral | 0.5 | 7 | 4.3 | 3.96 | 1.08 | 9.4 | 7.94 | 1.18 | Wu et al. 2017 |

Table S9: The input parameters for the development of ticagrelor PBPK model.

| Parameter | Value in the recent model | Unit | Reference |
| --- | --- | --- | --- |
| Molecular weight (MW) | 522.58 | g/mol | (7) |
| pKa (acid) | 12.94 | – | (7) |
| Solubility (water) | 0.063 | g/L | (7) |
| logP | 3.6 | – | (8) |
| Fraction unbound | 0.002 | - | (7) |
| CYP3A4 Km (AM metabolite) | 11 | µmol/L | (8) |
| CYP3A4 kcat (AM metabolite) | 170 | 1/min | Optimized |
| CYP3A4 Km (AR-C133913XX metabolite) | 41.3 | µmol/L | (8) |
| CYP3A4 kcat (AR-C133913XX metabolite) | 174 | 1/min | Optimized |
| Renal plasma clearance | 0.0584 | –L/h | (8) |
| Partition coefficients | Poulin and Theil | – | Optimized |
| Cellular permeability | 0.01 | cm/min | Optimized |
| Intestinal permeability | 2.9 × 10^-4^ | cm/min | Optimized |
| Particle dissolution thickness | 30 | µmol | Default |
| Particle dissolution particle radius | 10 | µmol | Default |

Table S10: Clinical studies used for ticagrelor PBPK model development and validation.

| Author year | N | Age (year) | Weight (kg) | Gender (% female) | Dose  (mg) | Route | Population |
| --- | --- | --- | --- | --- | --- | --- | --- |
| Holmberg et al., 2013(9) | 10 | 22 | 75 (57–95) | 40 | 90 | Oral | Young adult |
| Teng et al.,2014(10) | 12 | 32 | NR | 0 | 15 | IV | Young adult |
| Simona Rizea-Savu et al.,2025(11) | 28 | 37.7 | NR | 39.3 | 90 | Oral | Young adult |
| Teng et al.,2012 (Young, male)(12) | 10 | 29.0 | NR | 0 | 200 | Oral | Young adult |
| Teng et al.,2012 (Young, female)(12) | 9 | 40.3 | NR | 100 | 200 | Oral | Young adult |
| Holmberg et al.,2019(13) | 13 | 25 | 72 | 46 | 90 | Oral | Young adult |
| Teng et al.,2010(14) | 9 | 39 | 77 | 7.7 | 100 | Oral | Young adult |
| Teng et al.,2010(14) | 8 | 39 | 77 | 7.7 | 200 | Oral | Young adult |
| Teng et al.,2010(14) | 8 | 39 | 77 | 7.7 | 300 | Oral | Young adult |
| Teng et al.,2010(14) | 7 | 39 | 77 | 7.7 | 400 | Oral | Young adult |
| Butler et al.,2010(15) | 14 | 38 | 72.5 | 10 | 50 (BID) | Oral | Young adult |
| Butler et al.,2010(15) | 13 | 38 | 72.5 | 10 | 100 (BID) | Oral | Young adult |
| Butler et al.,2010(15) | 13 | 38 | 72.5 | 10 | 200 (BID) | Oral | Young adult |
| Butler et al.,2010(15) | 7 | 33 | 73 | 10 | 300 (BID) | Oral | Young adult |
| Butler et al.,2010(15) | 7 | 43 | 72 | 10 | 50 (QD) | Oral | Young adult |
| Butler et al.,2010(15) | 7 | 43 | 72 | 10 | 100 (QD) | Oral | Young adult |
| Butler et al.,2010(15) | 14 | 38 | 72.5 | 10 | 200 (QD) | Oral | Young adult |
| Butler et al.,2010(15) | 7 | 33 | 73 | 10 | 300 (QD) | Oral | Young adult |
| Butler et al.,2010(15) | 6 | 33 | 73 | 10 | 400 (QD) | Oral | Young adult |
| Butler et al.,2010(15) | 6 | 33 | 73 | 10 | 600 (QD) | Oral | Young adult |
| Teng et al.,2013(16) | 13 | 37 | 73 | 12 | 50 (BID) | Oral | Young adult |
| Teng et al.,2013(16) | 13 | 37 | 73 | 12 | 200 (BID) | Oral | Young adult |
| Teng et al.,2012 (12) | 10 | 67.5 | NR | 0 | 200 | Oral | Elderly |
| Teng et al.,2012(12) | 10 | 70.1 | NR | 100 | 200 | Oral | Elderly |

Table S11: Mean relative deviation (MRD) of ticagrelor plasma concentration prediction.

| Regimen | Route | MRD | Reference |
| --- | --- | --- | --- |
| 200 mg BID | Oral | 1.26 | Butler 2010 |
| 600 mg QD | Oral | 1.29 | Butler 2010 |
| 90 mg | Oral | 1.29 | Simona Rizea-Savu 2025 |
| 300 mg BID | Oral | 1.32 | Butler 2010 |
| 50 mg BID | Oral | 1.38 | Teng 2013 |
| 100 mg BID | Oral | 1.38 | Butler 2010 |
| 400 mg QD | Oral | 1.39 | Butler 2010 |
| 100 mg | Oral | 1.47 | Teng 2010 |
| 50 mg BID | Oral | 1.47 | Butler 2010 |
| 200 mg | Oral | 1.50 | Teng 2012, young male |
| 300 mg QD | Oral | 1.52 | Butler 2010 |
| 200 mg | Oral | 1.54 | Teng 2010 |
| 100 mg QD | Oral | 1.58 | Butler 2010 |
| 200 mg BID | Oral | 1.62 | Teng 2013 |
| 15 mg | Oral | 1.62 | Teng 2014 |
| 300 mg | Oral | 1.69 | Teng 2010 |
| 200 mg | Oral | 1.71 | Teng 2012, young female |
| 90 mg | Oral | 1.74 | Holmberg 2019 |
| 200 mg QD | Oral | 1.76 | Butler 2010 |
| 50 mg QD | Oral | 1.86 | Butler 2010 |
| 90 mg | Oral | 1.93 | Holmberg 2013 |
| 400 mg | Oral | 2.08 | Teng 2010 |
|  | Average | 1.56 |  |

Table S12: Predicted and observed ticagrelor AUClast and Cmax values with geometric mean fold errors (GMFEs) in base model.

| Author year | Dose  (mg) | Route | Predicted Cmax  (ng/ml) | Observed Cmax  (ng/ml) | GMEF Cmax | Predicted AUC  (ng*h/ml) | Observed AUC  (ng*h/ml) | GMFE AUC |
| --- | --- | --- | --- | --- | --- | --- | --- | --- |
| Holmberg 2013 | 90 | Oral | 520 | 497 | 1.05 | 4084 | 2745 | 1.49 |
| Teng 2014 | 15 | IV | 618 | 449 | 1.38 | 1148 | 1058 | 1.09 |
| Simona Rizea-Savu 2025 | 90 | Oral | 455 | 492 | 1.08 | 4118 | 2804 | 1.47 |
| Simona Rizea-Savu 2025 | 90 | Oral | 455 | 549 | 1.21 | 4118 | 3060 | 1.35 |
| Teng 2012 (Young, male) | 200 | Oral | 955 | 672 | 1.42 | 9403 | 5753 | 1.63 |
| Teng 2012 (Young, female) | 200 | Oral | 1004 | 1080 | 1.08 | 10907 | 8819 | 1.24 |
| Holmberg 2019 | 90 | Oral | 477 | 408 | 1.17 | 4286 | 2150 | 1.99 |
| Teng 2010 | 100 | Oral | 493 | 586 | 1.19 | 4568 | 3683 | 1.24 |
| Teng 2010 | 200 | Oral | 919 | 1295 | 1.41 | 9257 | 8213 | 1.13 |
| Teng 2010 | 300 | Oral | 1298 | 1746 | 1.35 | 12212 | 13170 | 1.08 |
| Teng 2010 | 400 | Oral | 1638 | 2711 | 1.66 | 16620 | 18547 | 1.12 |
| Butler 2010 | 50 (BID) | Oral | 348 | 251 | 1.39 | 2260 | 1682 | 1.34 |
| Butler 2010 | 100 (BID) | Oral | 714 | 626 | 1.14 | 4573 | 4108 | 1.11 |
| Butler 2010 | 200 (BID) | Oral | 1360 | 1440 | 1.06 | 8367 | 9436 | 1.13 |
| Butler 2010 | 300 (BID) | Oral | 1830 | 1936 | 1.06 | 11128 | 14203 | 1.28 |
| Butler 2010 | 50 (QD) | Oral | 297 | 222 | 1.34 | 2.295 | 1877 | 1.22 |
| Butler 2010 | 100 (QD) | Oral | 573 | 557 | 1.03 | 4645 | 4295 | 1.08 |
| Butler 2010 | 200 (QD) | Oral | 1101 | 1030 | 1.07 | 9299 | 7884 | 1.18 |
| Butler 2010 | 300 (QD) | Oral | 1598 | 1354 | 1.18 | 13279 | 10611 | 1.25 |
| Butler 2010 | 400 (QD) | Oral | 2003 | 1862 | 1.08 | 15833 | 14963 | 1.06 |
| Butler 2010 | 600 (QD) | Oral | 2662 | 2958 | 1.11 | 19998 | 24088 | 1.20 |
| Teng 2013 | 50 (BID) | Oral | 365 | 353 | 1.03 | 2446 | 2218 | 1.10 |
| Teng 2013 | 200 (BID) | Oral | 1336 | 1777 | 1.33 | 8616 | 12026 | 1.40 |
|  |  |  |  | Average Cmax GMFE | 1.21 | Average AUC GMFE | 1.28 |  |

Table S13: Mean relative deviation (MRD) of ticagrelor plasma concentration prediction in geriatric population.

| Regimen | Route | MRD | Reference |
| --- | --- | --- | --- |
| 200 mg | Oral | 1.39 | Teng 2012, Old male |
| 200 mg | Oral | 1.69 | Teng 2012, old female |
|  | Average MRD | 1.54 |  |

Table S14: Predicted and observed rosuvastatin AUClast and Cmax values with geometric mean fold errors (GMFEs) in geriatric population.

| Author year | Dose  (mg) | Route | Predicted Cmax  (ng/ml) | Observed Cmax  (ng/ml) | GMEF Cmax | Predicted AUC  (ng*h/ml) | Observed AUC  (ng*h/ml) | GMFE AUC |
| --- | --- | --- | --- | --- | --- | --- | --- | --- |
| Teng 2012 (Old, male) | 200 | Oral | 1110 | 1160 | 1.05 | 14962 | 9752 | 1.53 |
| Teng 2012 (Old, female) | 200 | Oral | 1188 | 1666 | 1.40 | 16859 | 11965 | 1.41 |
|  |  |  |  | Average GMEF Cmax | 1.22 |  | Average GMEF AUC | 1.47 |

Table S15: Predicted and Observed Ticagrelor-Rosuvastatin DDI Ratios for AUC_last_ and C_max_ with Geometric Mean Fold Errors (GMFEs) in subjects with wild type genotype of *ABCG2* c.421C>A.

| *ABCG2* c.421C>A | Perpetrator | Victim | Dose gap [h] | n | DDI AUC _last_ ratio | | | DDI C_max_ ratio | | | Reference |
| --- | --- | --- | --- | --- | --- | --- | --- | --- | --- | --- | --- |
|  |  |  |  |  | Pred | Obs | GMFE | Pred | Obs | GMFE |  |
| Wildtype | Ticagrelor | 20.0 mg, po, tab | 1 | 9 | 2.25 | 2.47 | 1.09 | 1.92 | 2.58 | 1.34 | Lehtisalo et al. 2024 (4) |

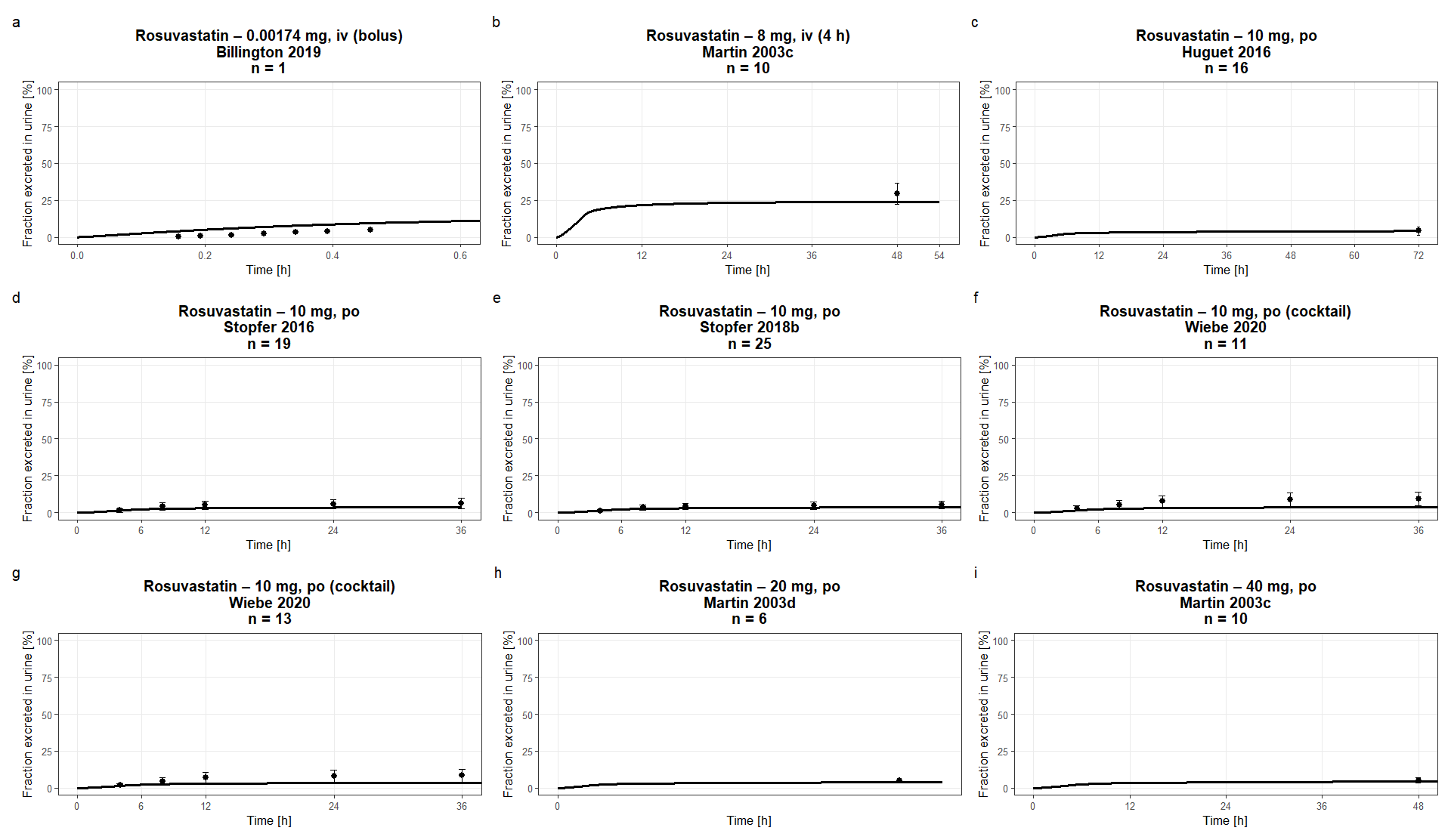

FigS1: Predicted versus observed fraction of rosuvastatin excreted in urine. Simulations are shown as lines, observed data are shown as dots. Details on administration protocols, study populations, and literature references are listed in previous study(1)

|  | A) | 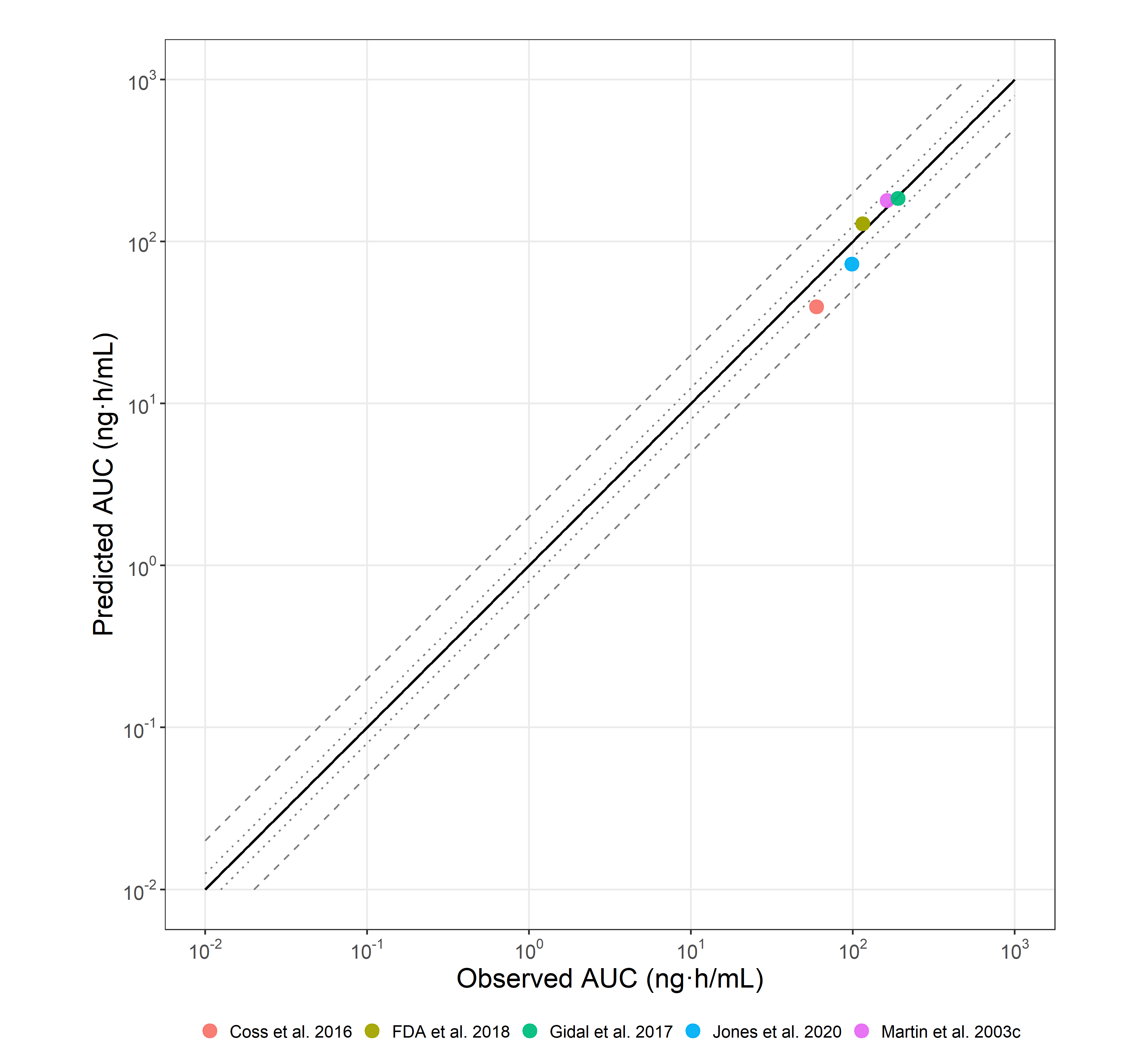 |
| --- | --- | --- |
|  | **B)** | 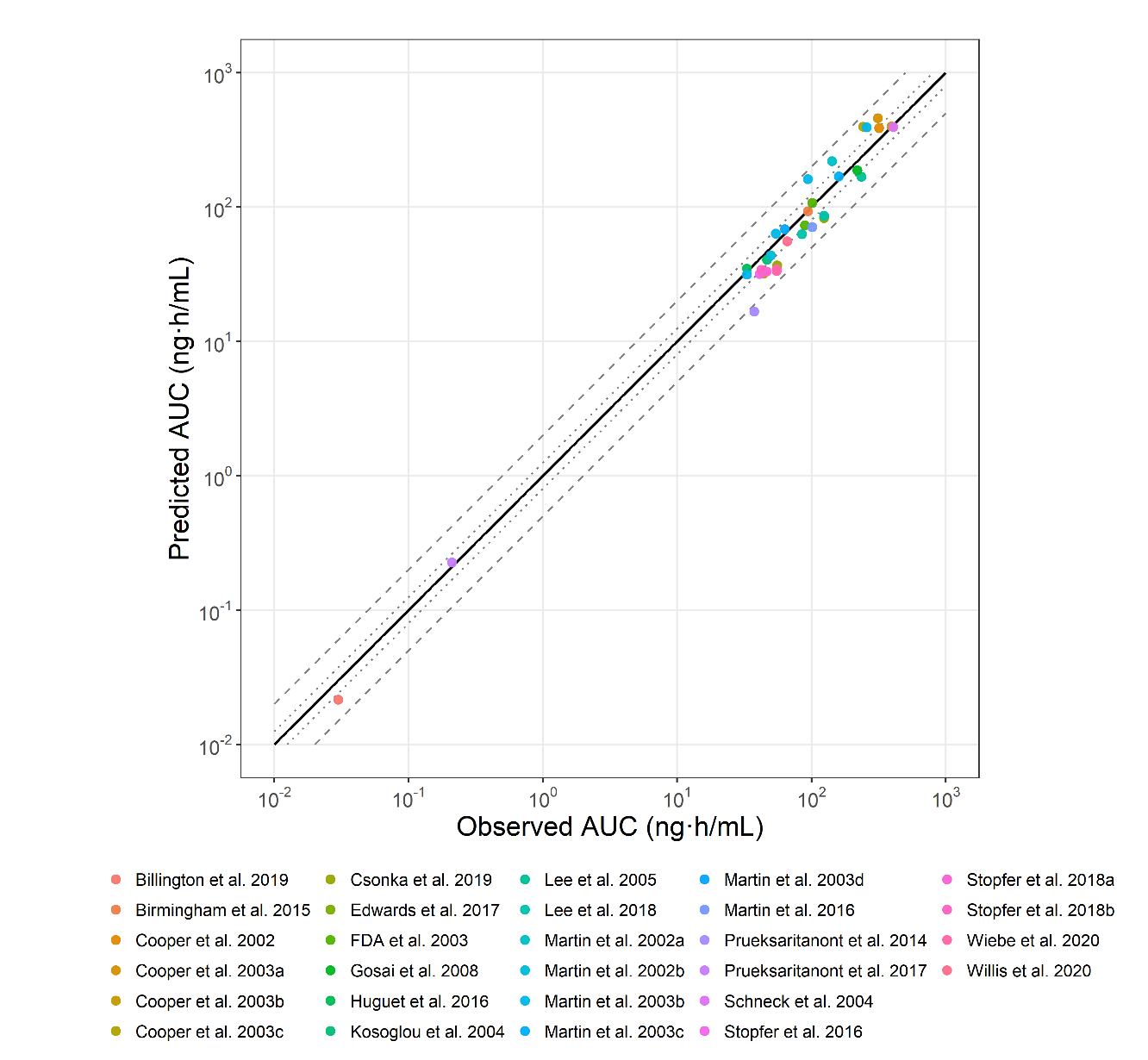 |
|  | FigS2: Predicted versus observed rosuvastatin AUClast values for the training (a) and test (b) datasets in the base model. Solid lines represent the line of identity. Dotted lines indicate 1.25-fold deviation; dashed lines indicate 2-fold deviation. Details on administration protocols, study populations, and literature references are listed in previous study(1). | |
| A) | | 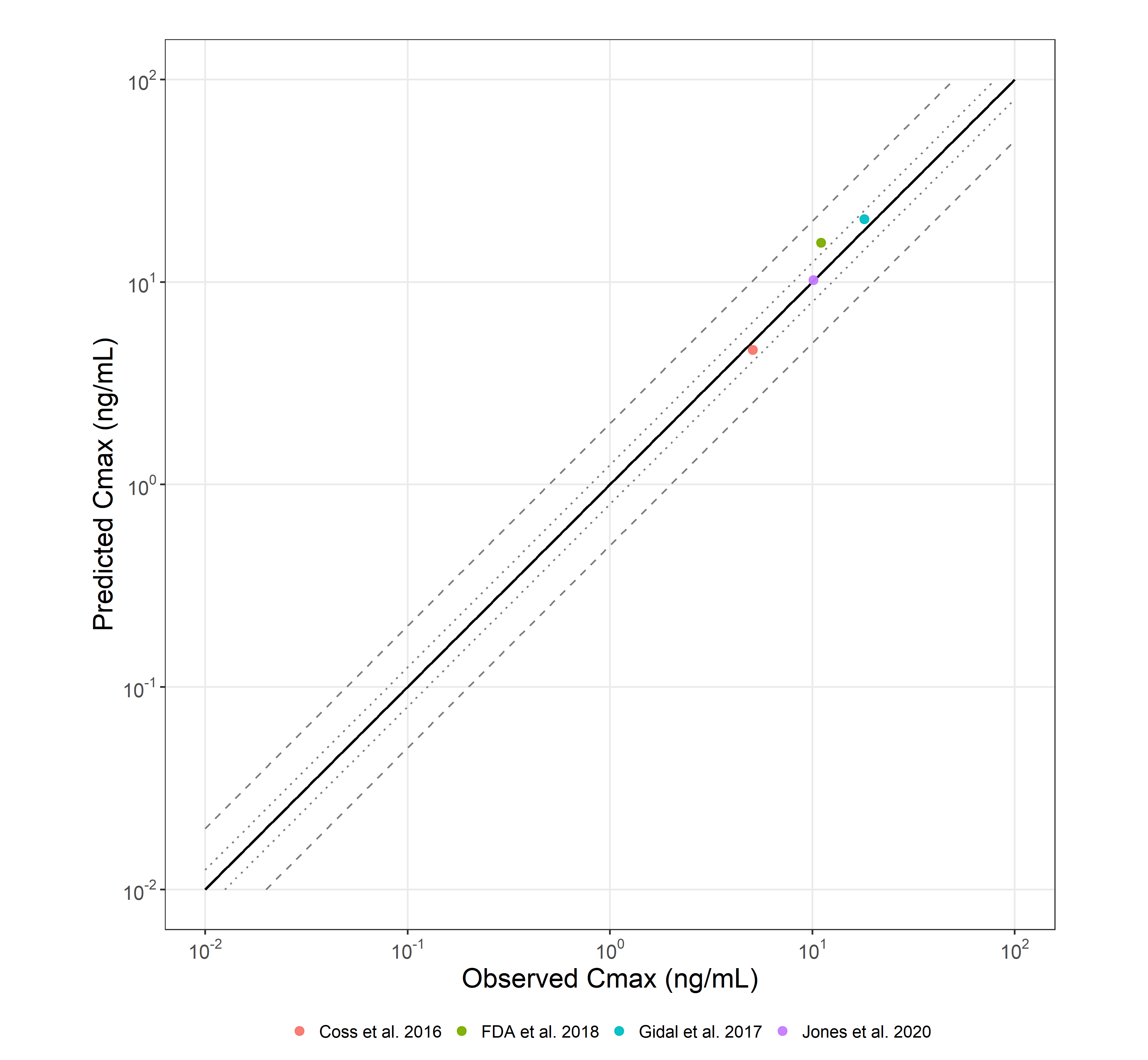 |
| B) | | 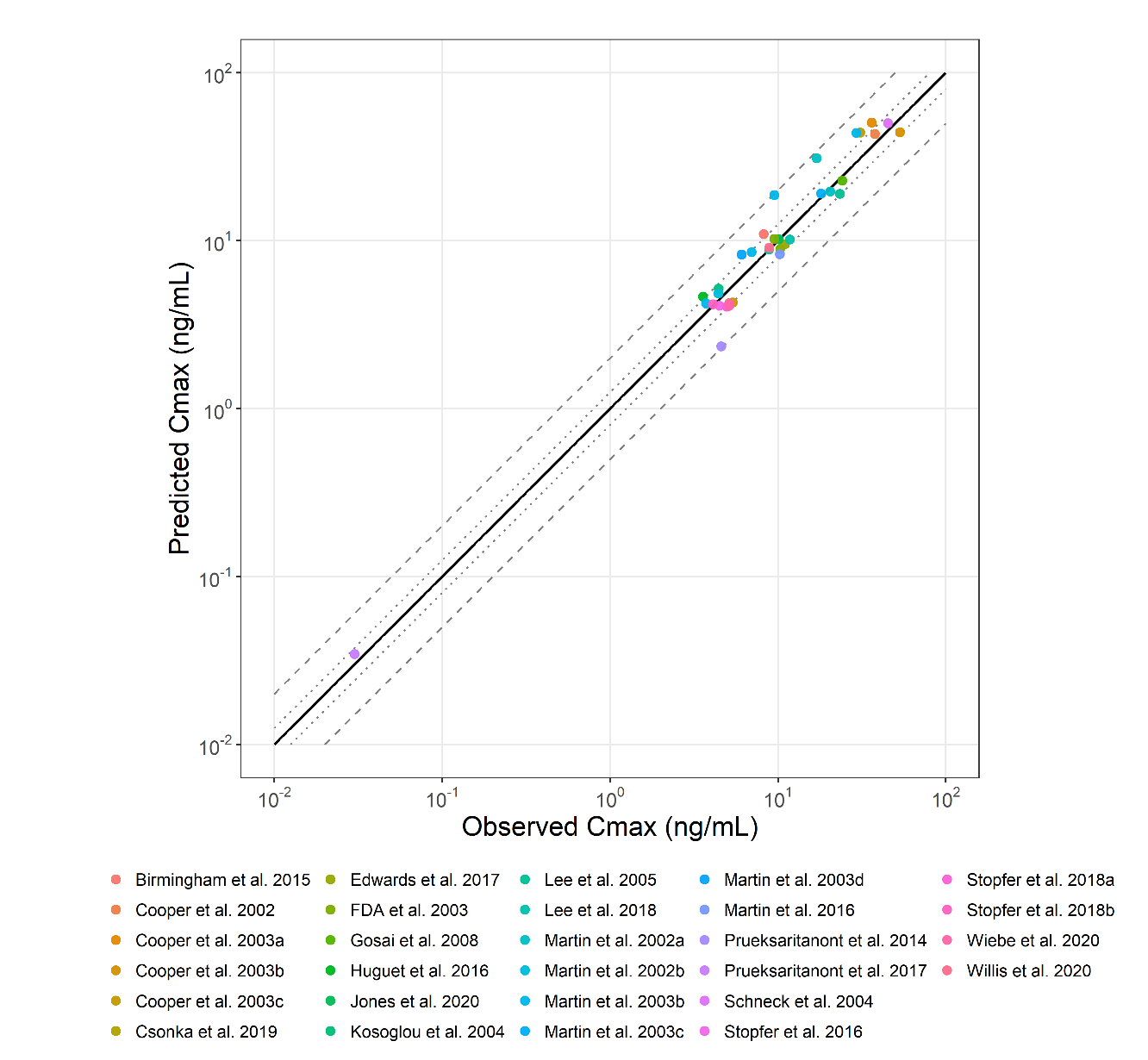 |
| FigS3: Predicted versus observed rosuvastatin Cmax values for the training (a) and test (b) datasets in the base model. Solid lines represent the line of identity. Dotted lines indicate 1.25-fold deviation; dashed lines indicate 2-fold deviation. Details on administration protocols, study populations, and literature references are listed in previous study(1) | | |

| A) | 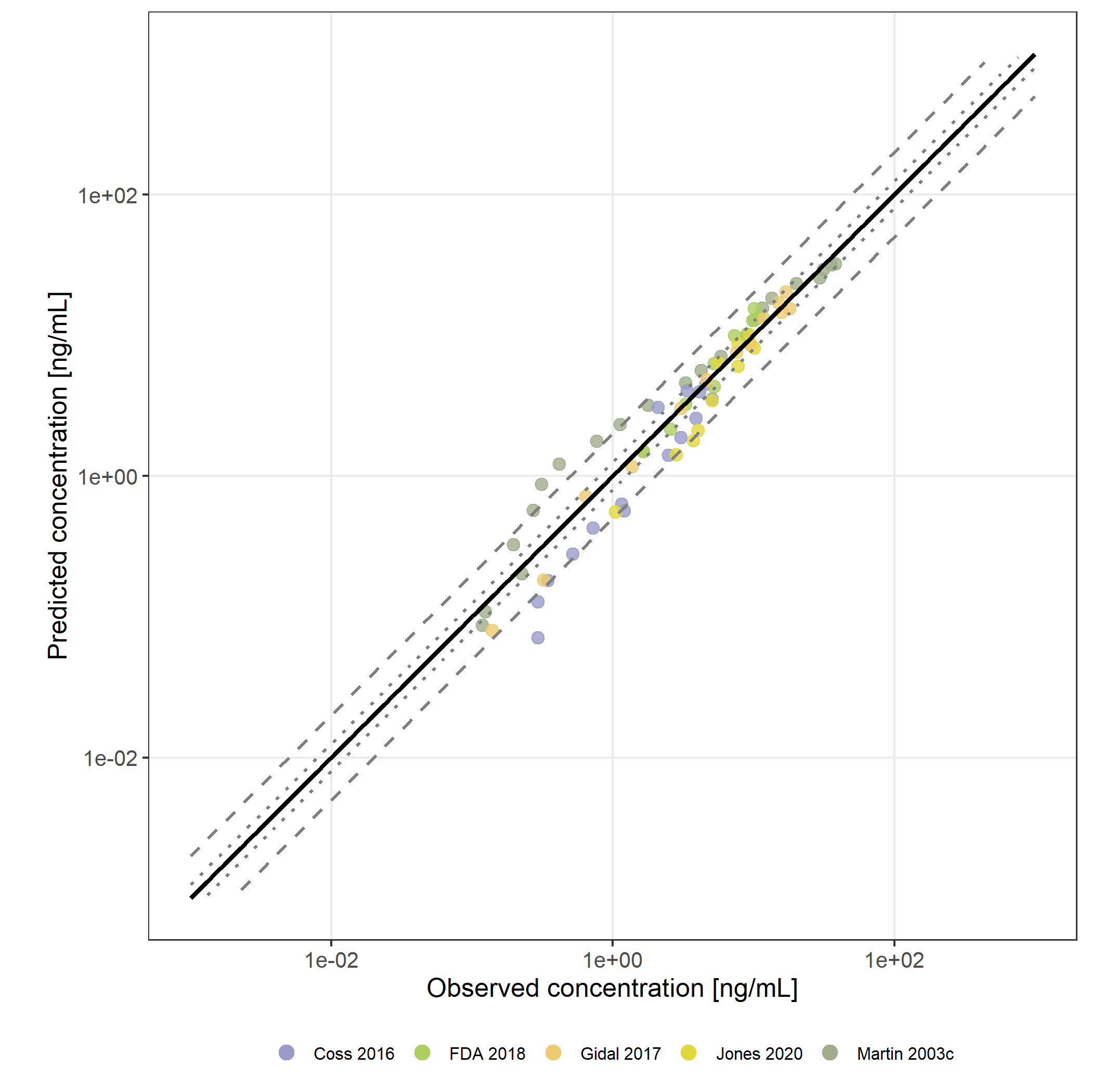 |
| --- | --- |
| B) | 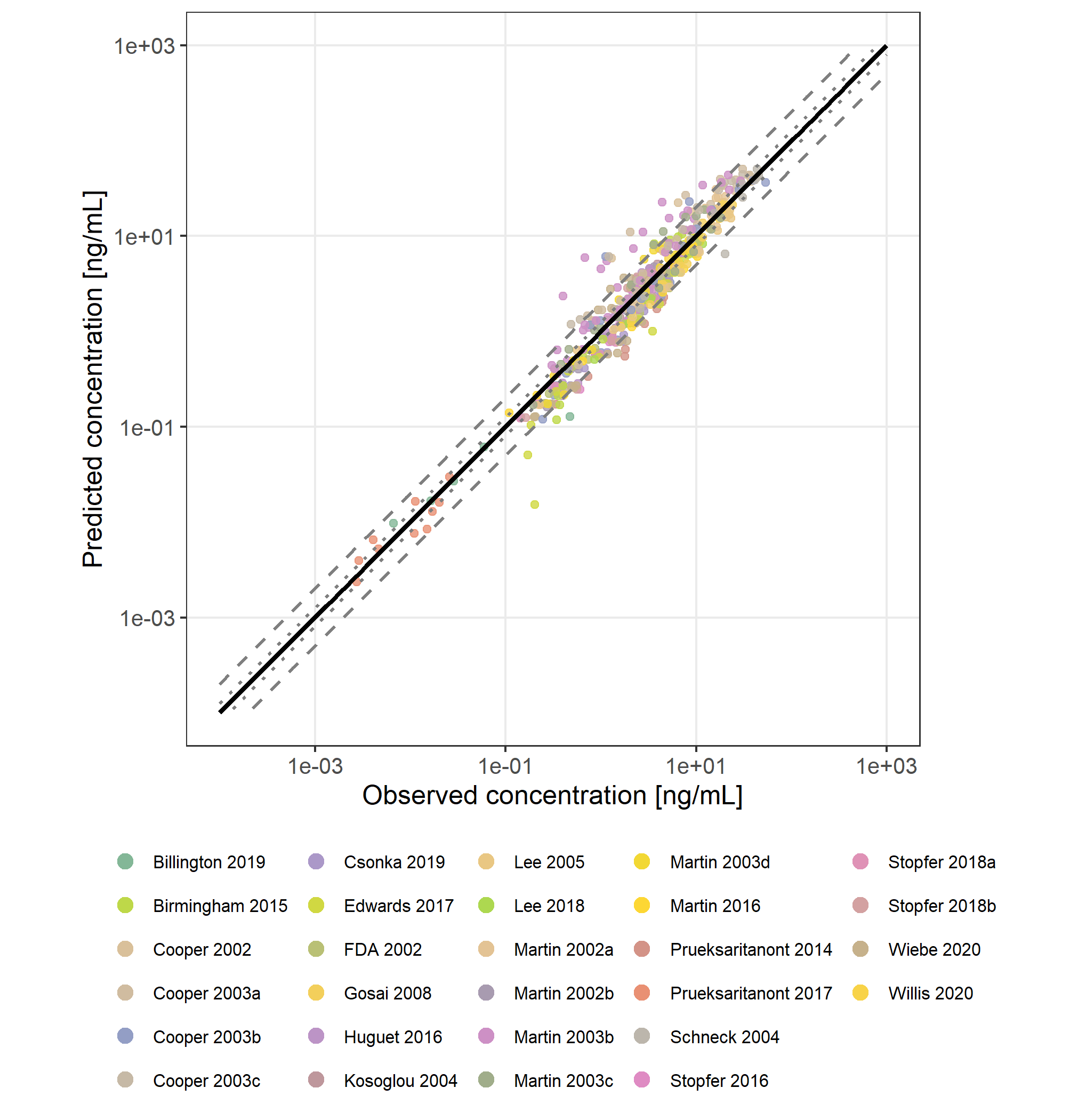 |
| FigS4: Predicted versus observed rosuvastatin plasma concentration values for the training (a) and test (b) datasets in the base model. Solid lines represent the line of identity. Dotted lines indicate 1.25-fold deviation; dashed lines indicate 2-fold deviation. Details on administration protocols, study populations, and literature references are listed in previous study(1) | |

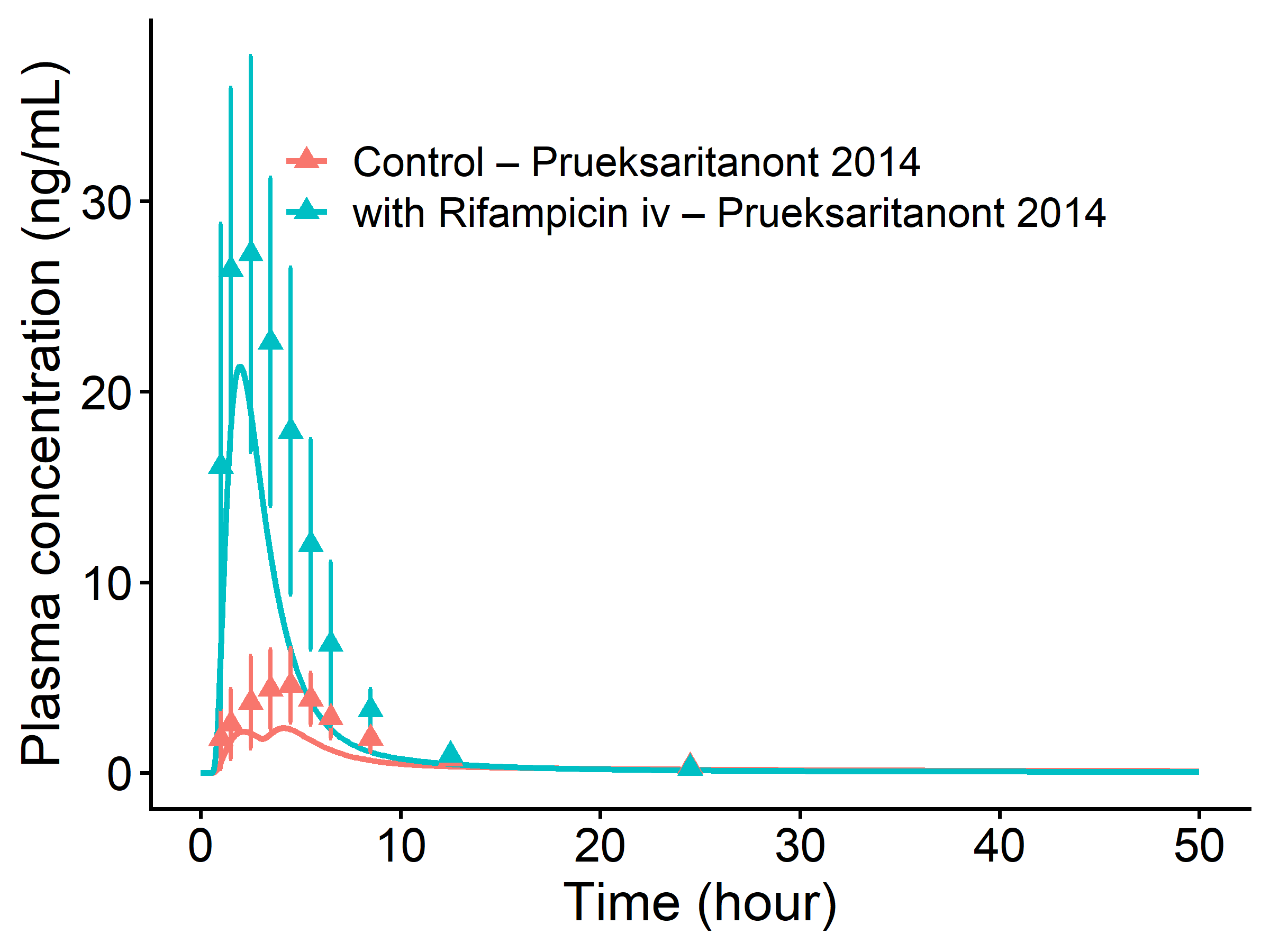

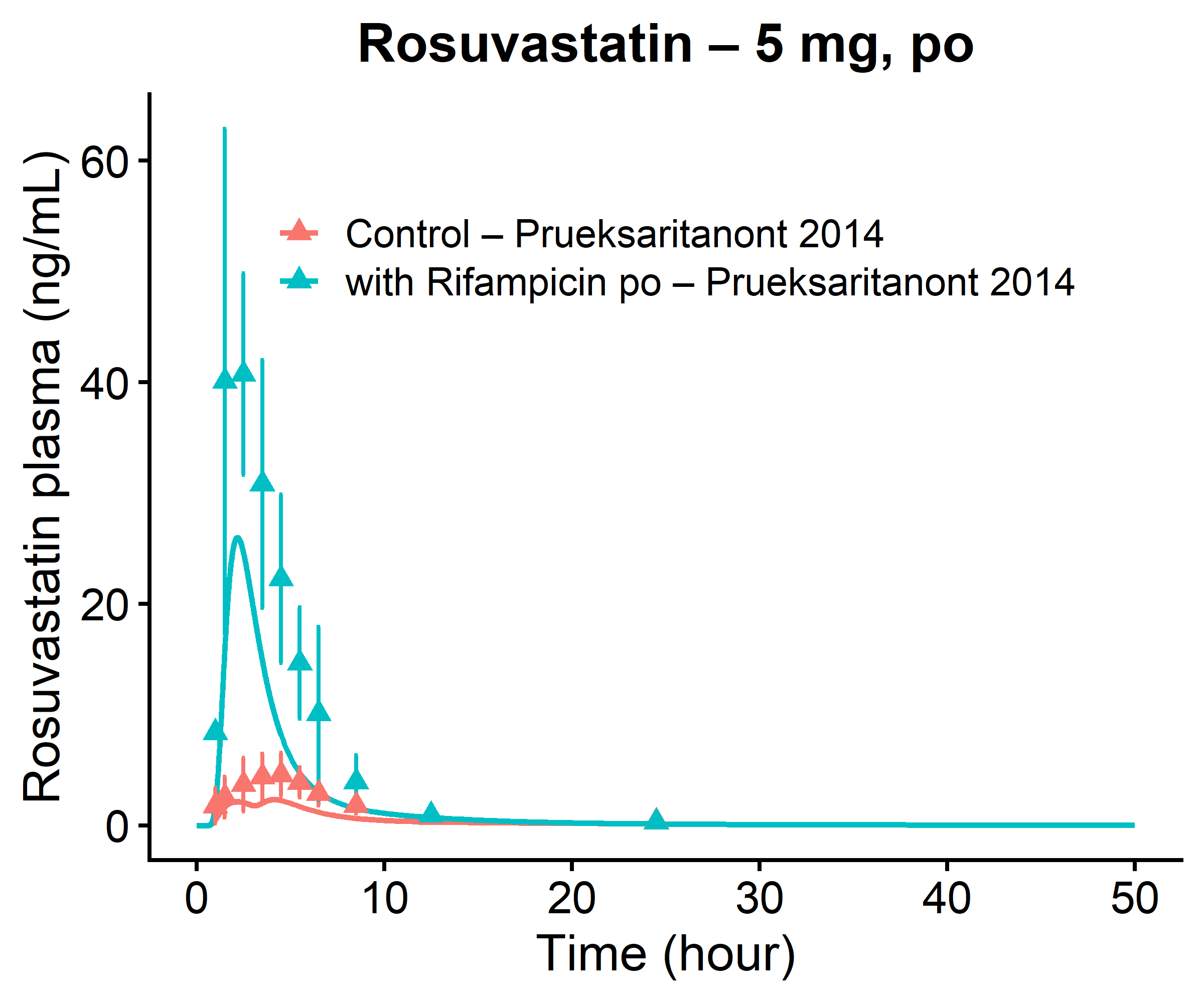

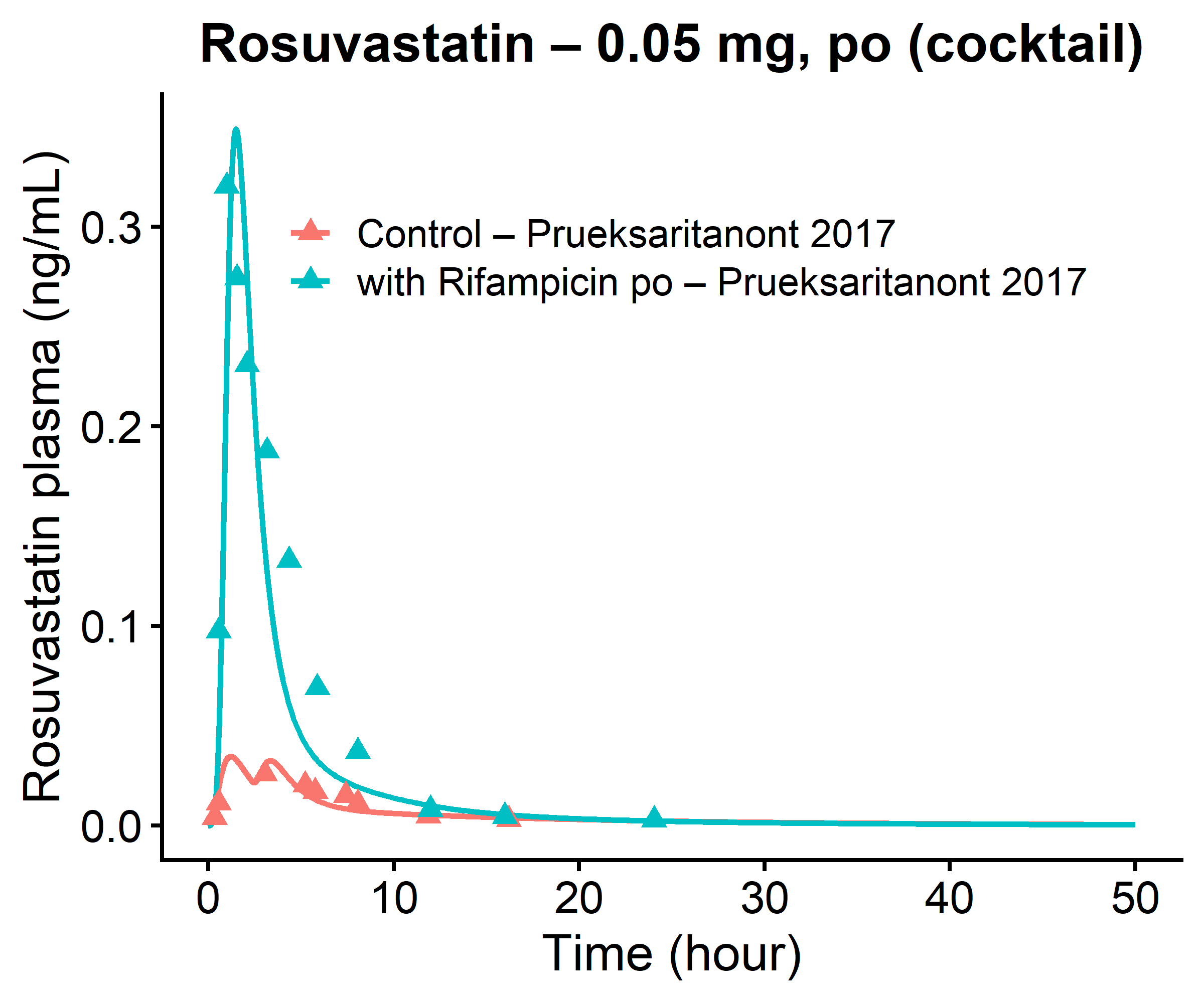

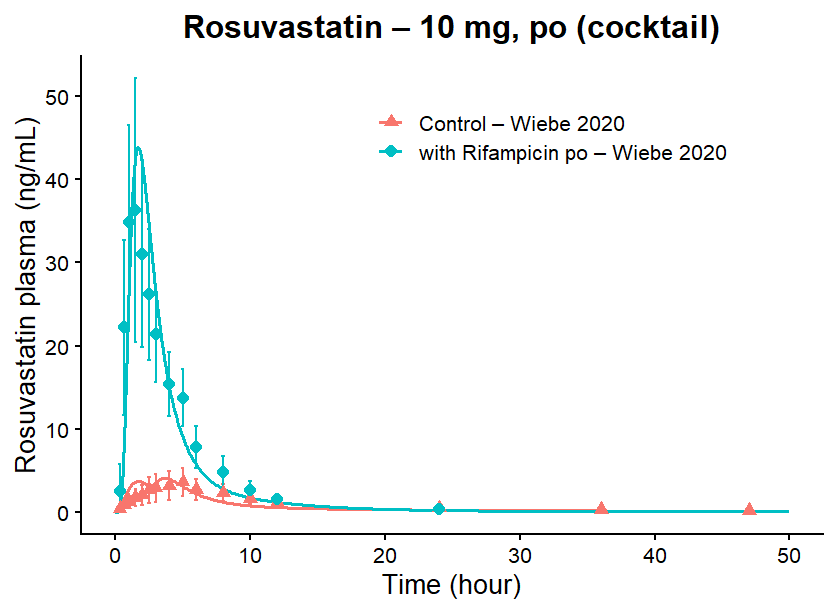

FigS5: Rosuvastatin plasma concentration-time profiles before and during rifampicin coadministration.Simulations are shown as lines, observed data are shown as dots. Details on administration protocols, study populations, and literature references are listed in previous study(1)

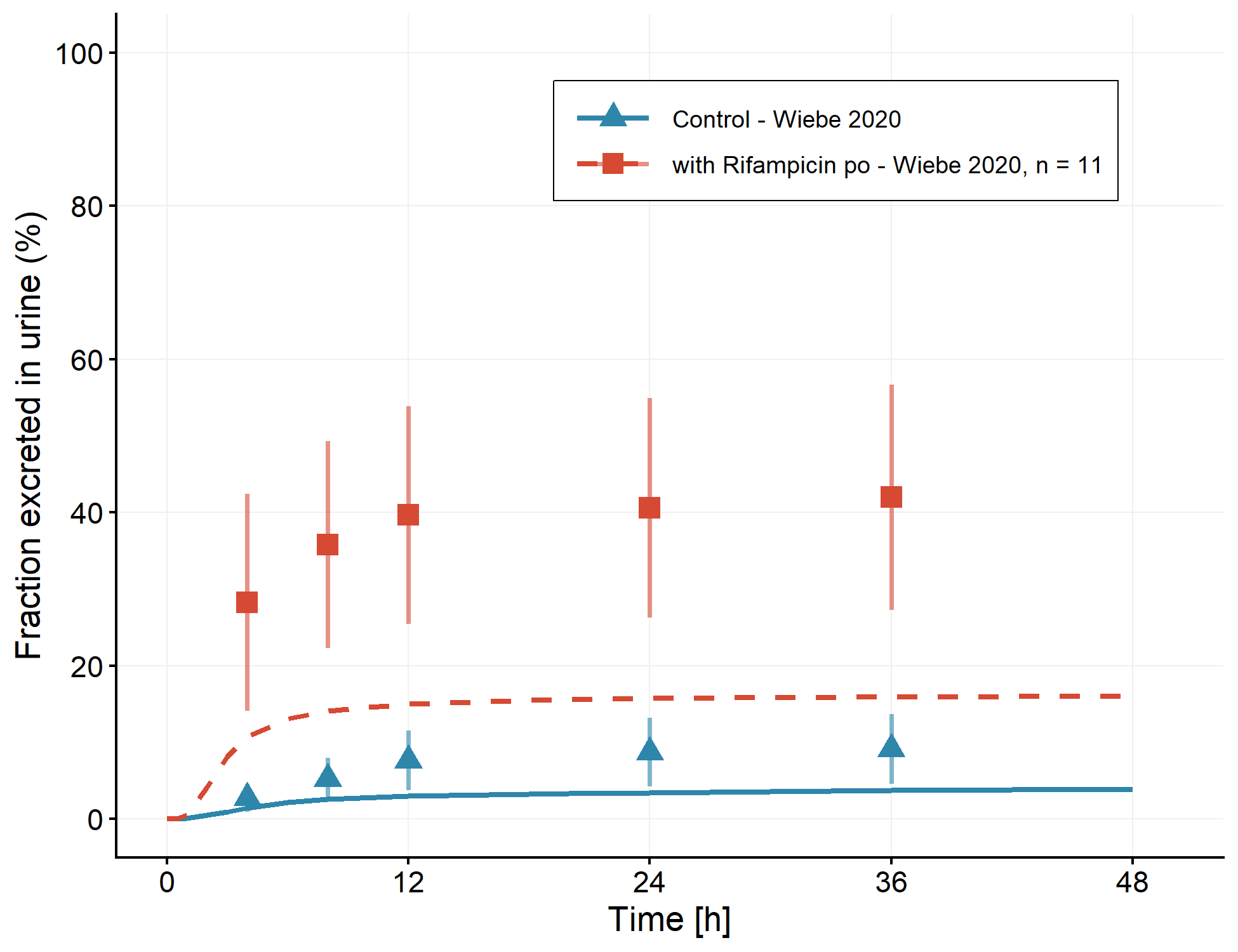

FigS6: Rosuvastatin fraction excreted in urine profiles, before and during oral rifampicin co-administration. Simulations are shown as lines, observed data are shown as dots. Details on administration protocols, study populations, and literature references are listed in previous study(1)

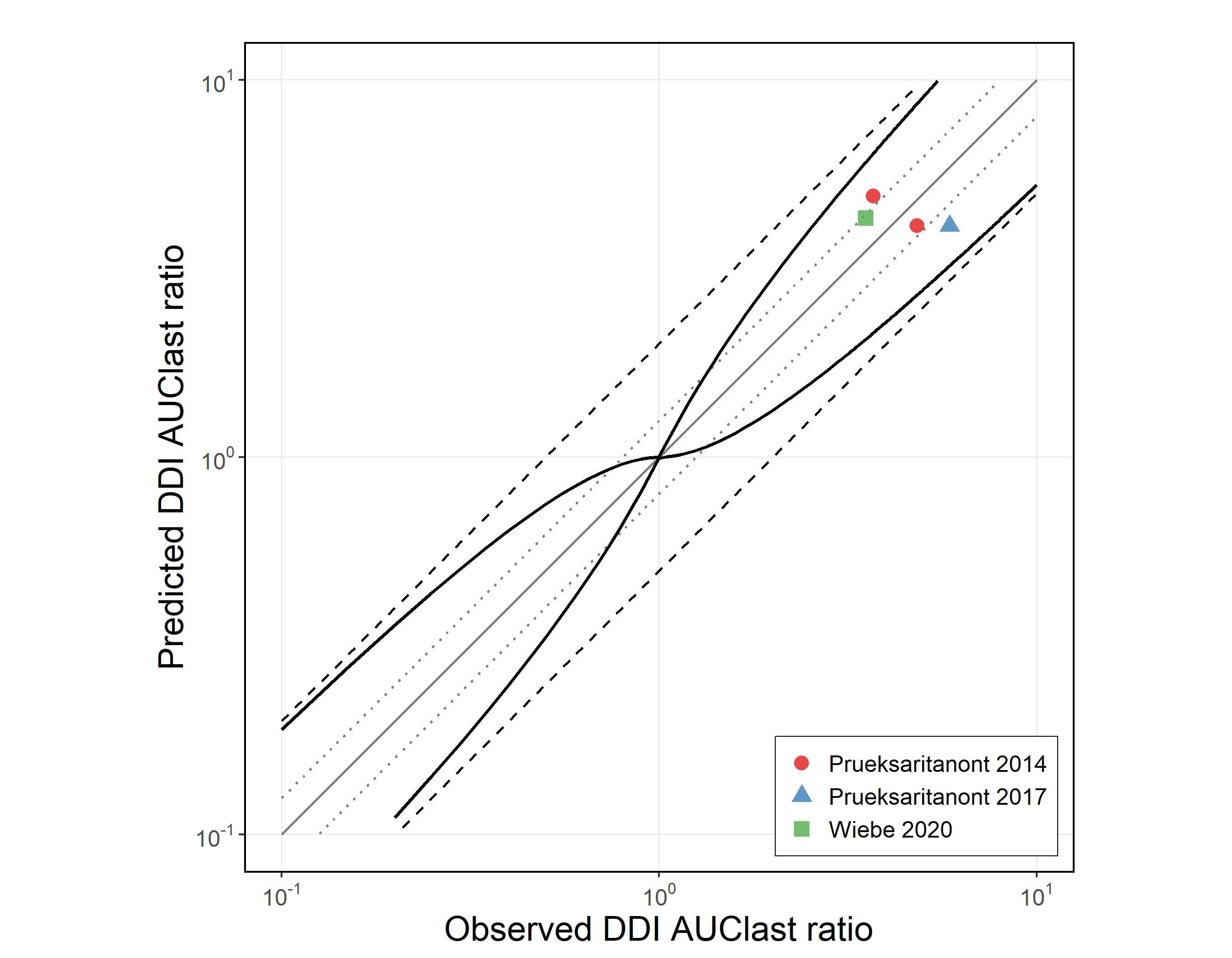

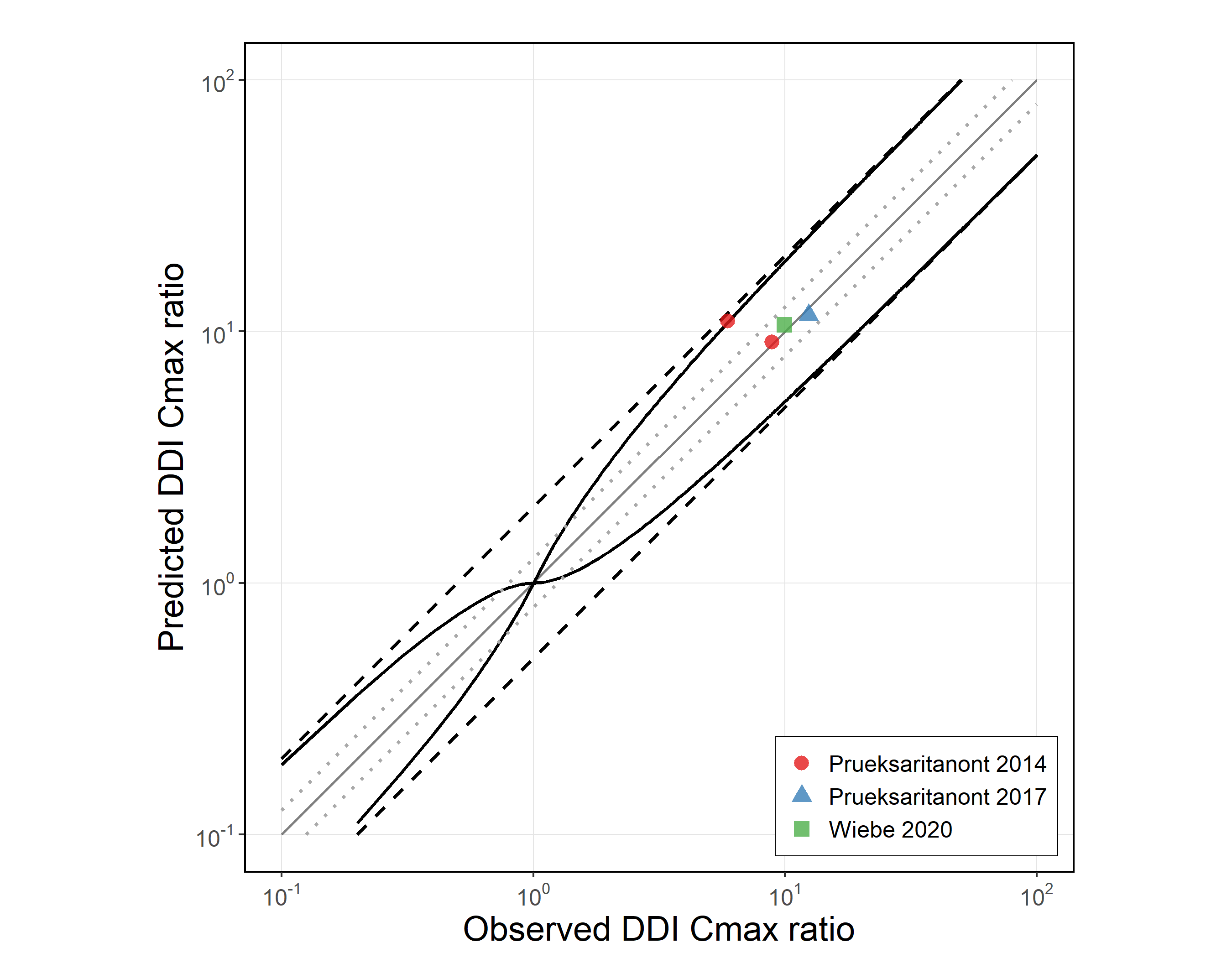

FigS7: Predicted versus observed rifampicin-rosuvastatin DDI AUClast ratios and DDI Cmax ratios in base model. The straight solid lines ( ) mark the line of identity. Dotted lines ( ) indicate 1.25-fold, dashed lines ( ) indicate 2-fold deviation. The curved solid lines illustrate the DDI prediction success limits suggested by Guest et al(2).

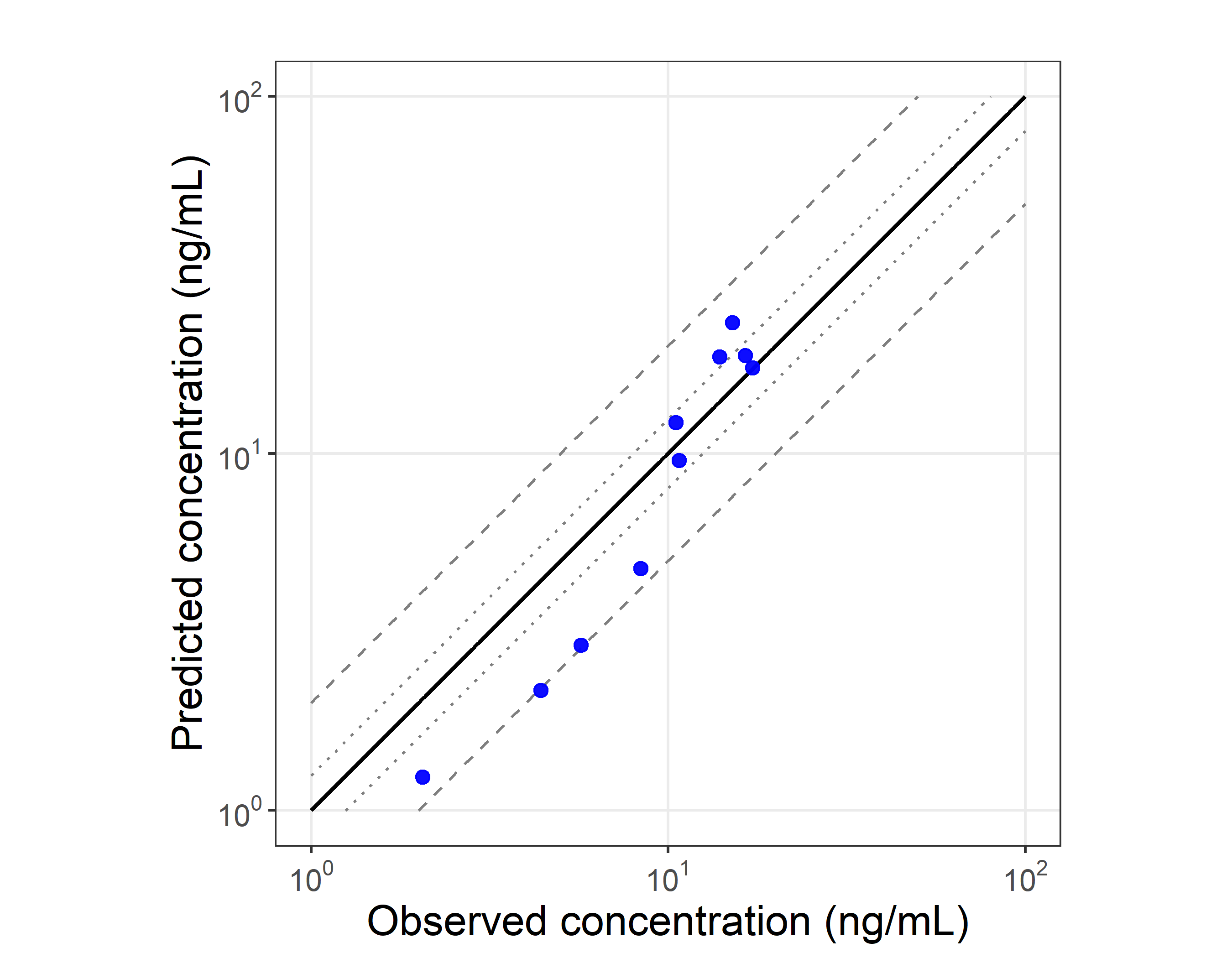

FigS8: Predicted versus observed rosuvastatin plasma concentration in model for geriatric population. Solid lines represent the line of identity. Dotted lines indicate 1.25-fold deviation; dashed lines indicate 2-fold deviation. Details on administration protocols, study populations, and literature references are listed in table S5.

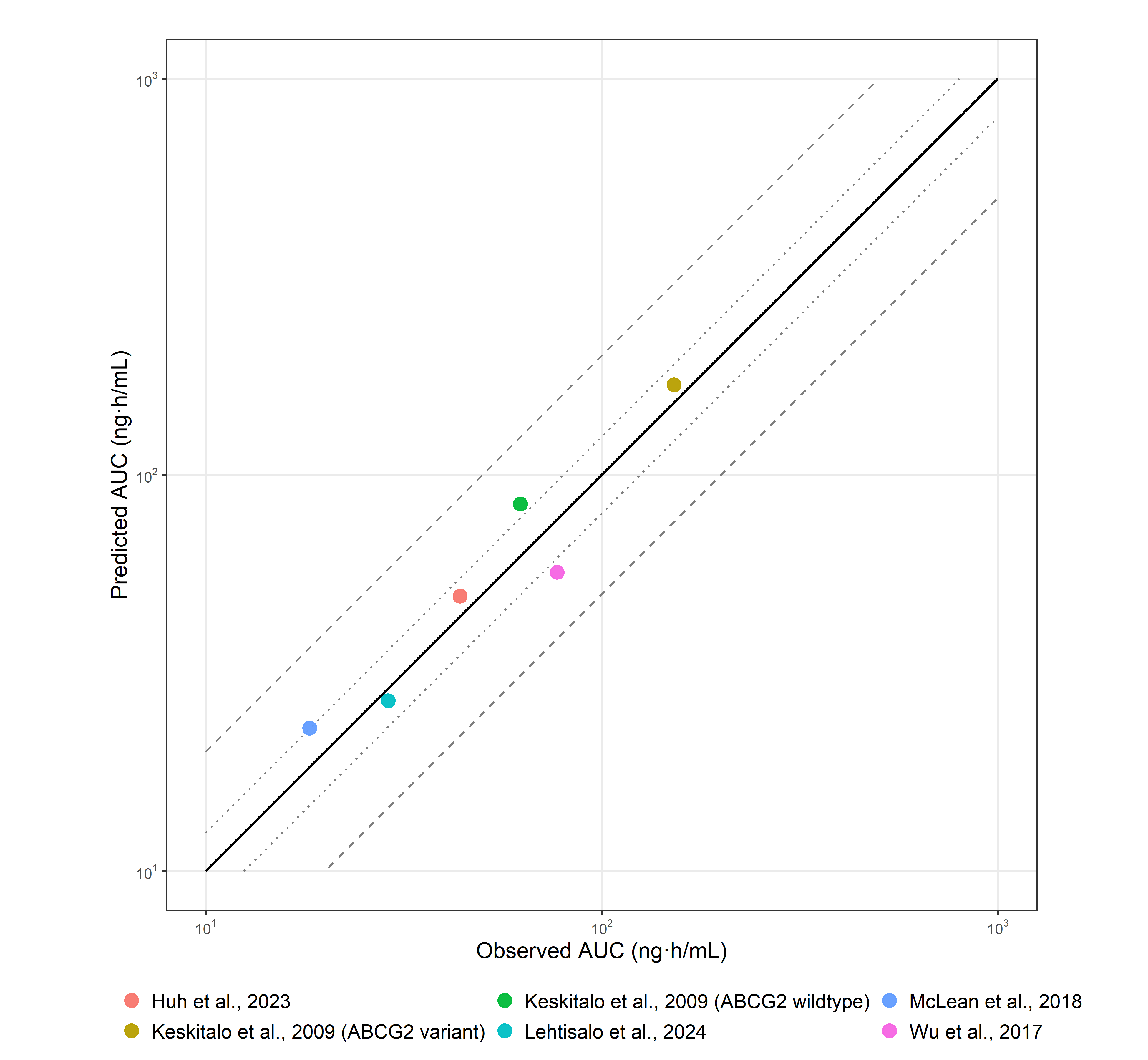

FigS9: Predicted versus observed rosuvastatin AUClast values for the *ABCG2* c.421C>A wild-type and variant (Keskito 2009) datasets in genetic model. Solid lines represent the line of identity. Dotted lines indicate 1.25-fold deviation; dashed lines indicate 2-fold deviation. Details on administration protocols, study populations, and literature references are listed in table S5.

|  | 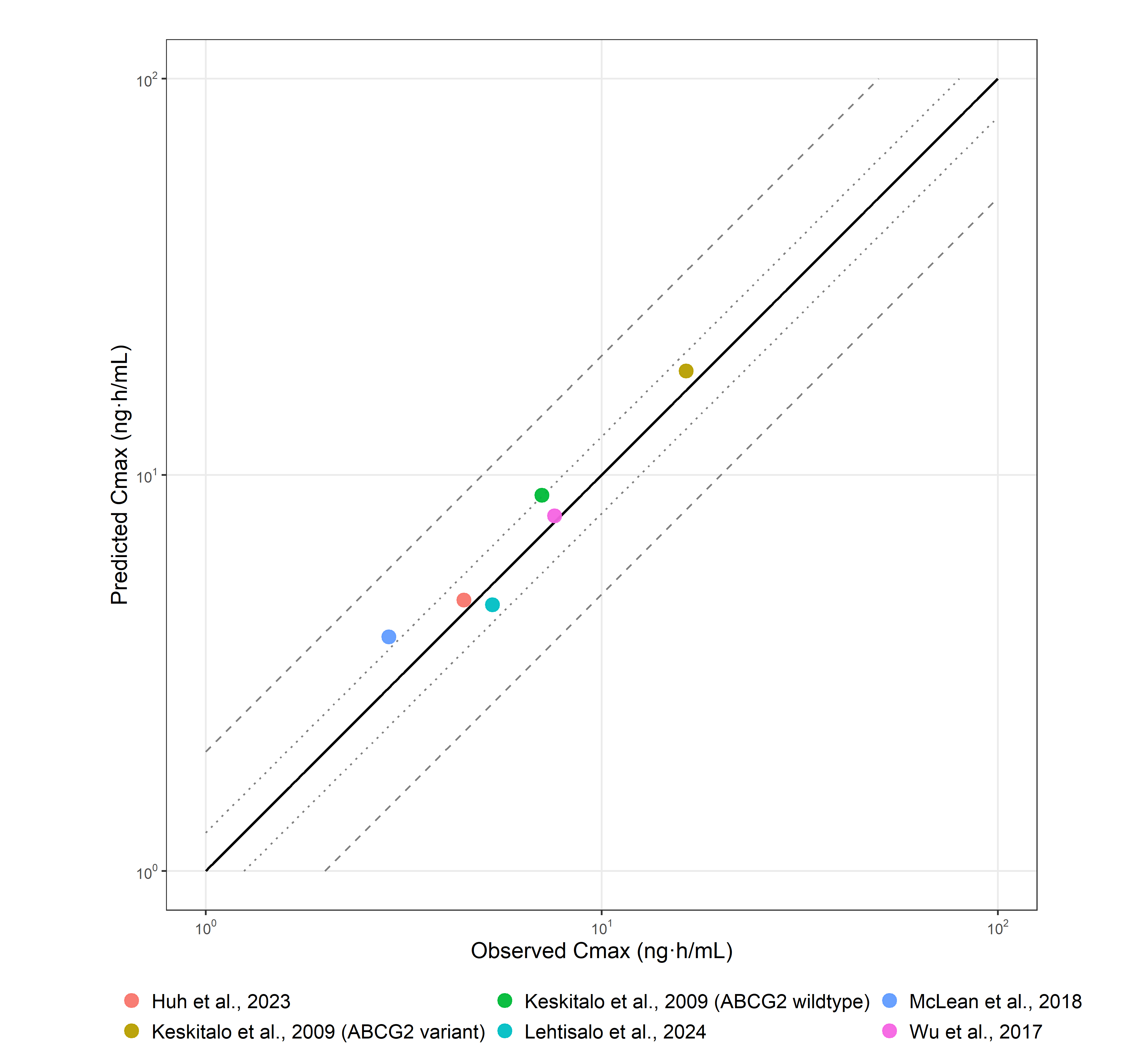 |
| --- | --- |
| FigS10: Predicted versus observed rosuvastatin Cmax values for the *ABCG2* c.421C>A wild-type and variant (Keskito 2009) datasets in genetic model. Solid lines represent the line of identity. Dotted lines indicate 1.25-fold deviation; dashed lines indicate 2-fold deviation. Details on administration protocols, study populations, and literature references are listed in table S5. | |

|  | 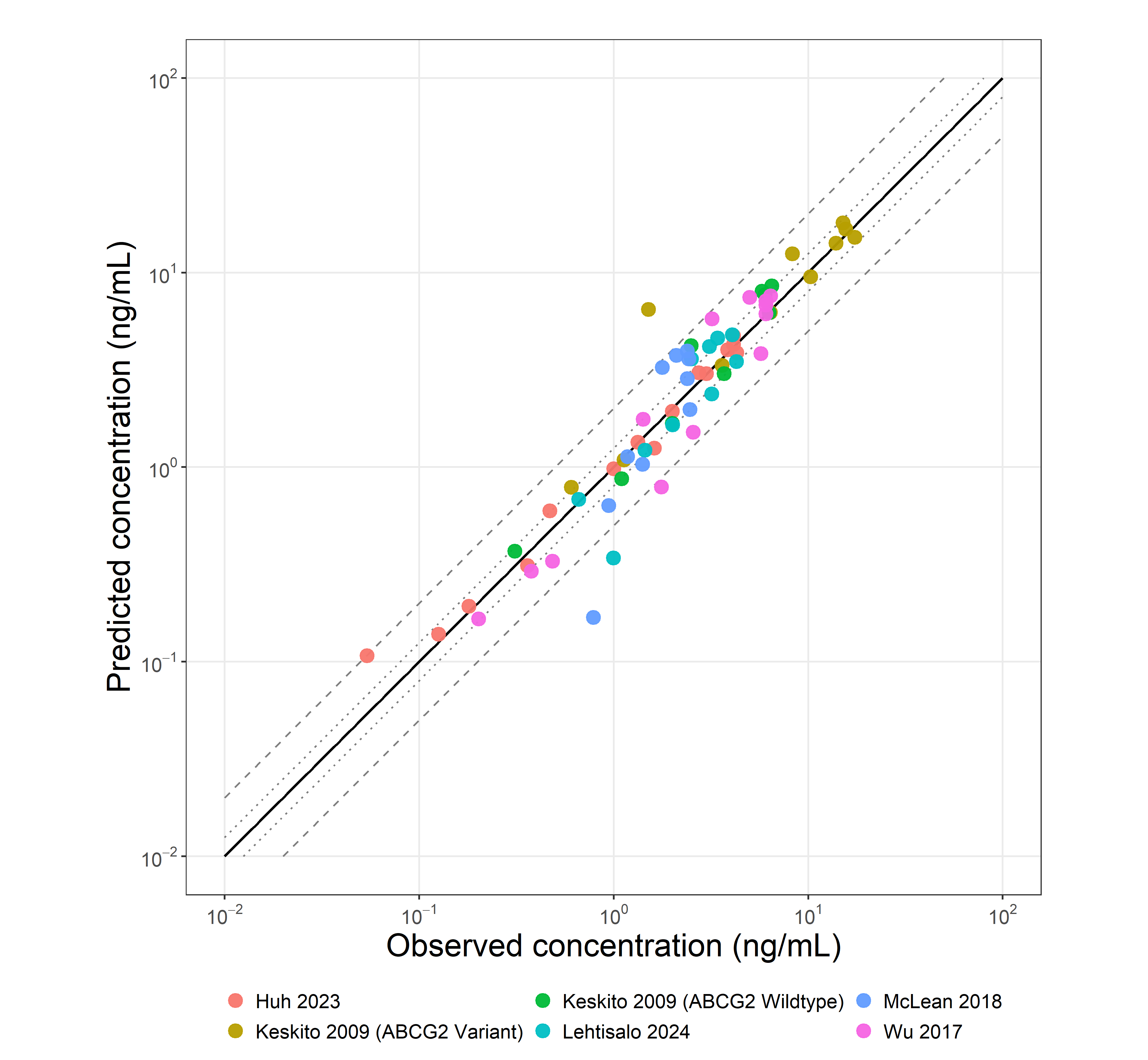 |
| --- | --- |
| FigS11: Predicted versus observed rosuvastatin plasma concentration values for the *ABCG2* c.421C>A wild-type and variant (Keskito 2009) datasets in genetic model. Solid lines represent the line of identity. Dotted lines indicate 1.25-fold deviation; dashed lines indicate 2-fold deviation. Details on administration protocols, study populations, and literature references are listed in table S5. | |

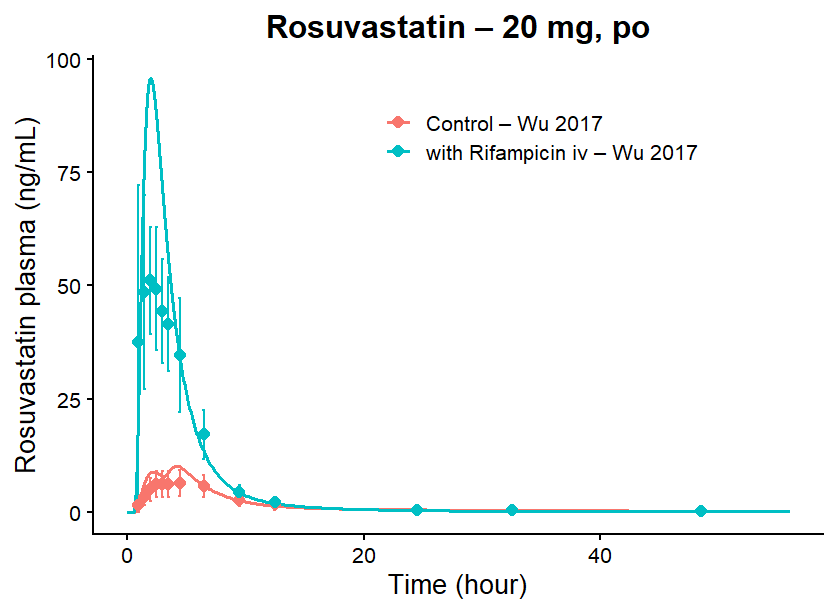

FigS12: Rosuvastatin plasma concentration-time profiles, before and during rifampicin coadministration in subjects with *ABCG2* c.421C>A wild-type genotype. Simulations are shown as lines, observed data are shown as dots.

| A) | 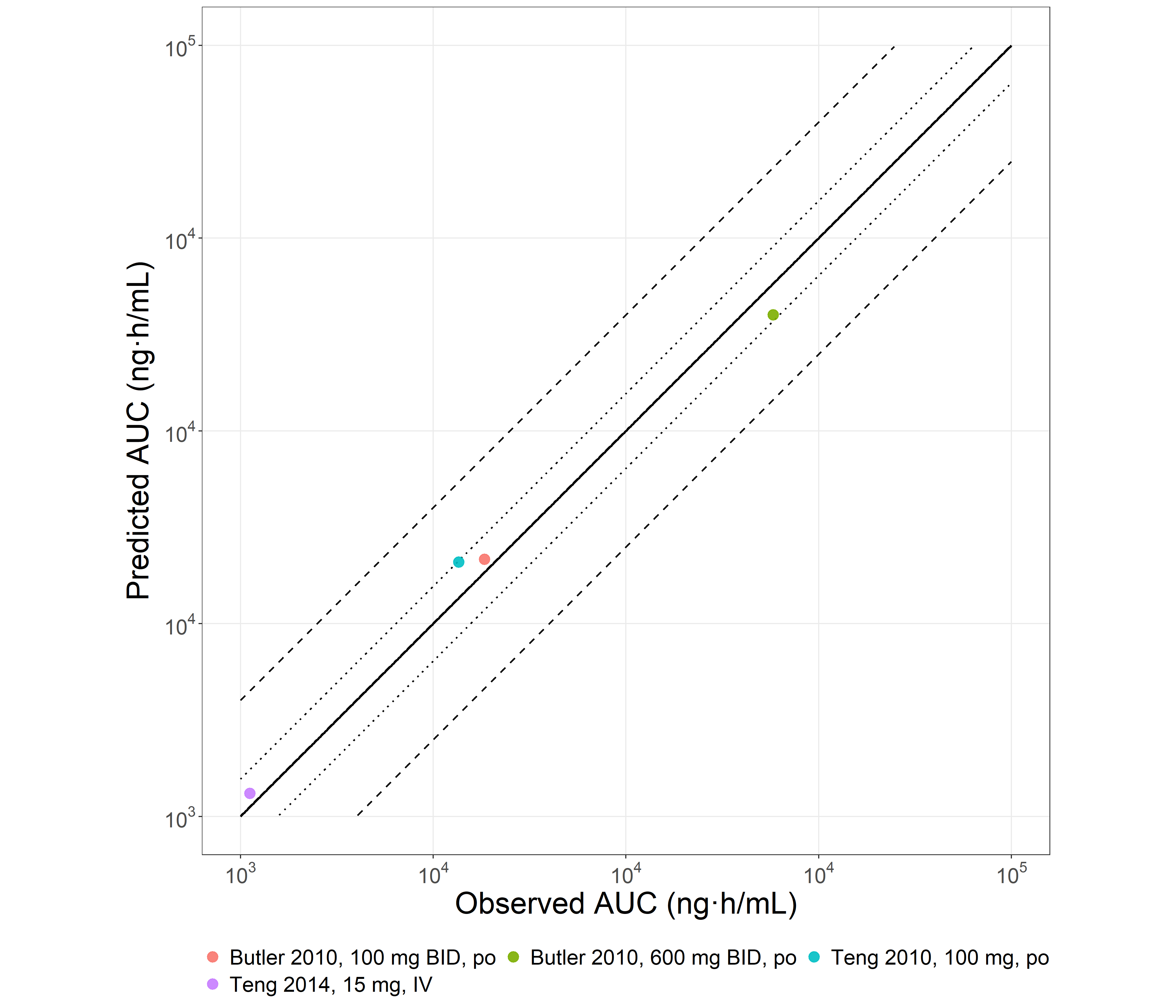 |
| --- | --- |
| B) | 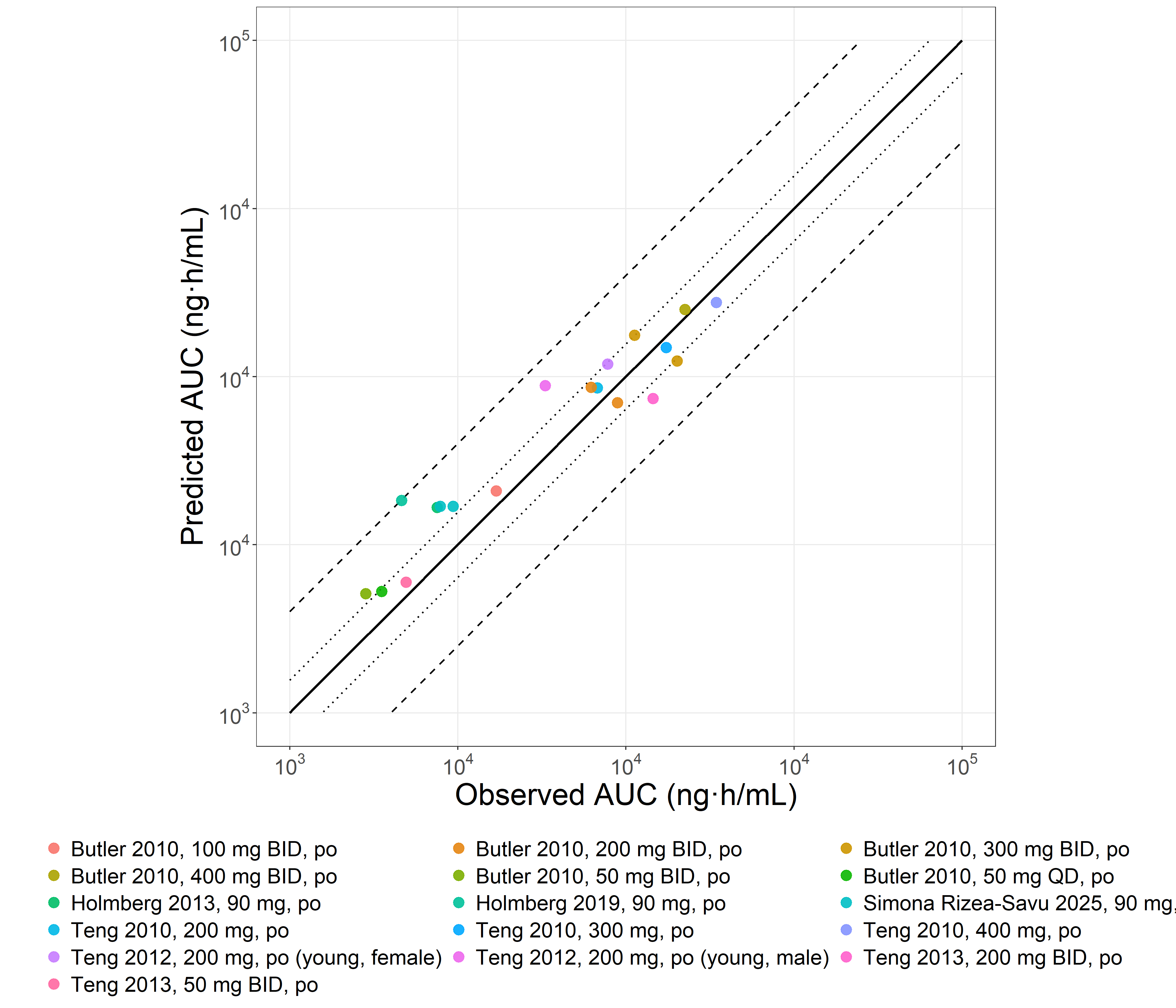 |
| FigS13: Predicted versus observed ticagrelor AUClast values for the training (a) and test (b) datasets in the base model. Solid lines represent the line of identity. Dotted lines indicate 1.25-fold deviation; dashed lines indicate 2-fold deviation. Details on administration protocols, study populations, and literature references are listed in table S10. | |

| A) | | 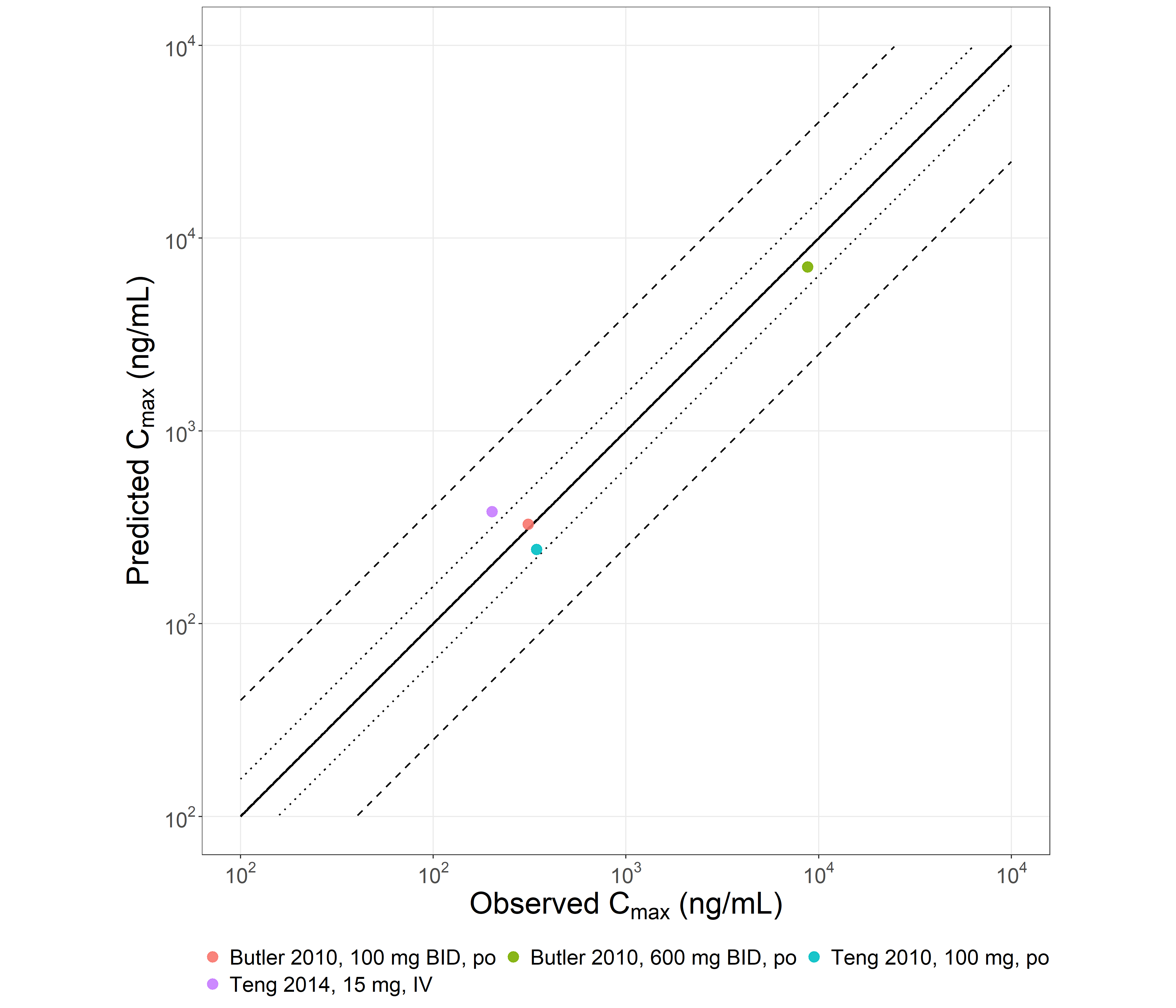 |
| --- | --- | --- |
| B) | 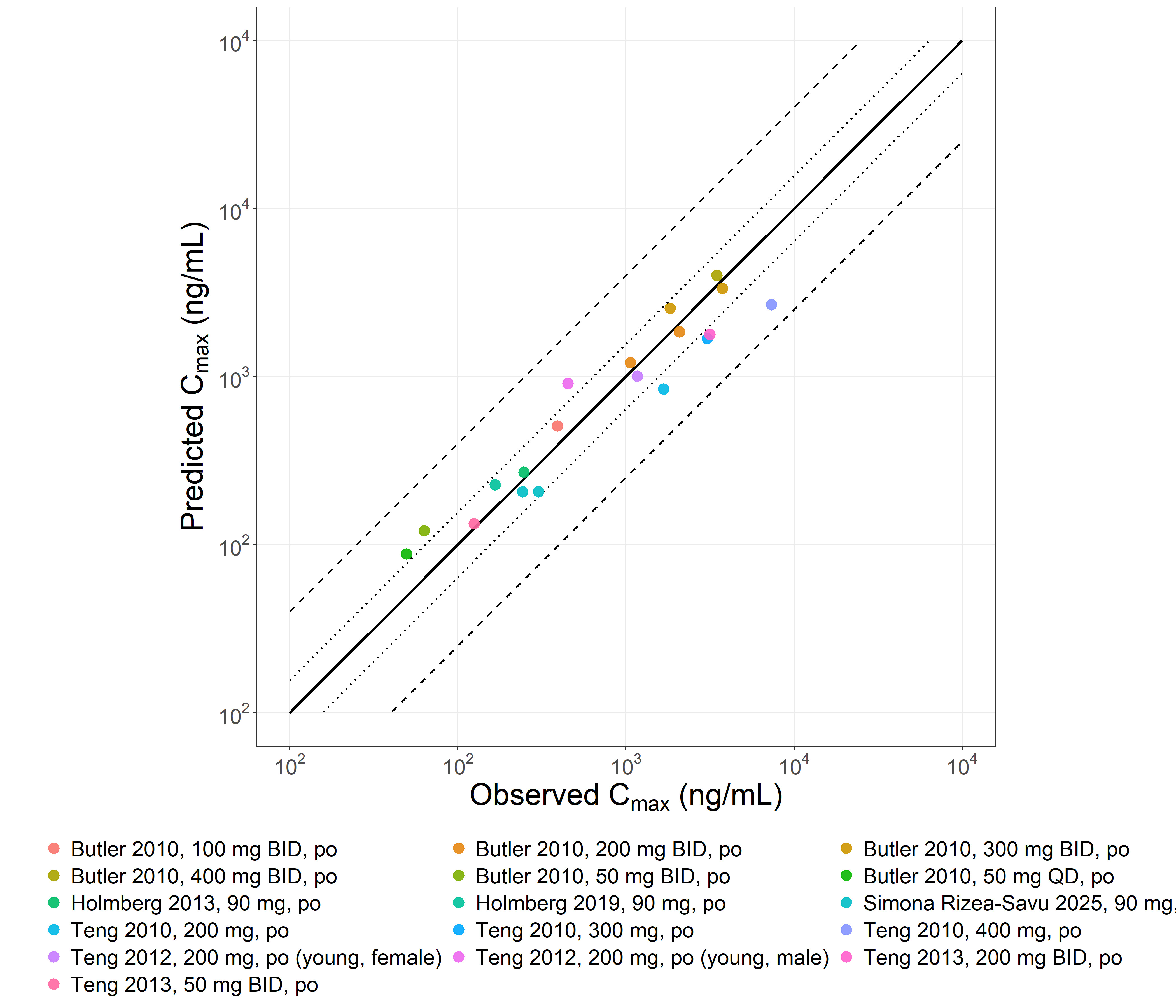 | |
| FigS14: Predicted versus observed ticagrelor Cmax values for the training (a) and test (b) datasets in the base model. Solid lines represent the line of identity. Dotted lines indicate 1.25-fold deviation; dashed lines indicate 2-fold deviation. Details on administration protocols, study populations, and literature references are listed in table S10. | | |

| A) | 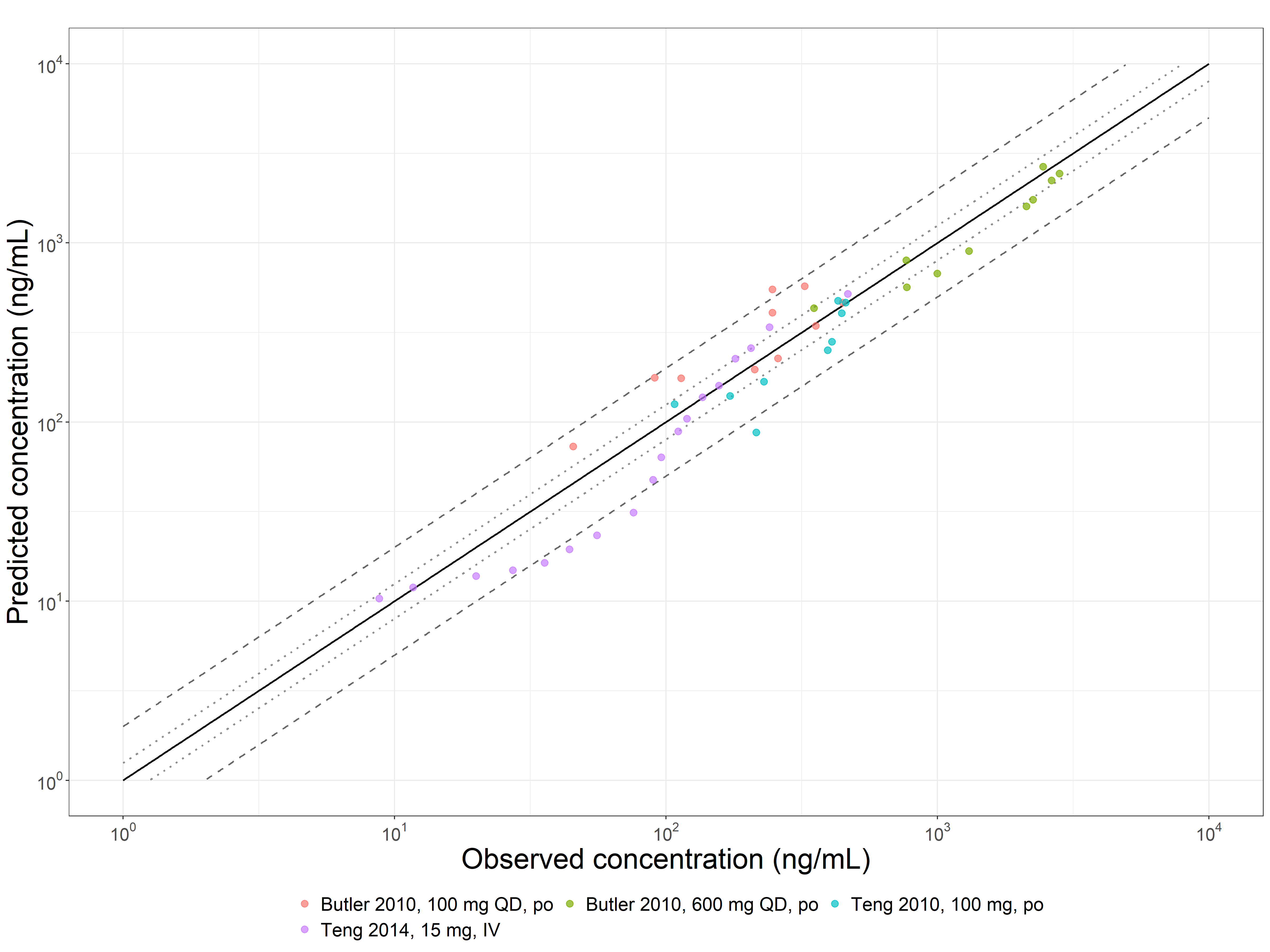 |
| --- | --- |
| B) | 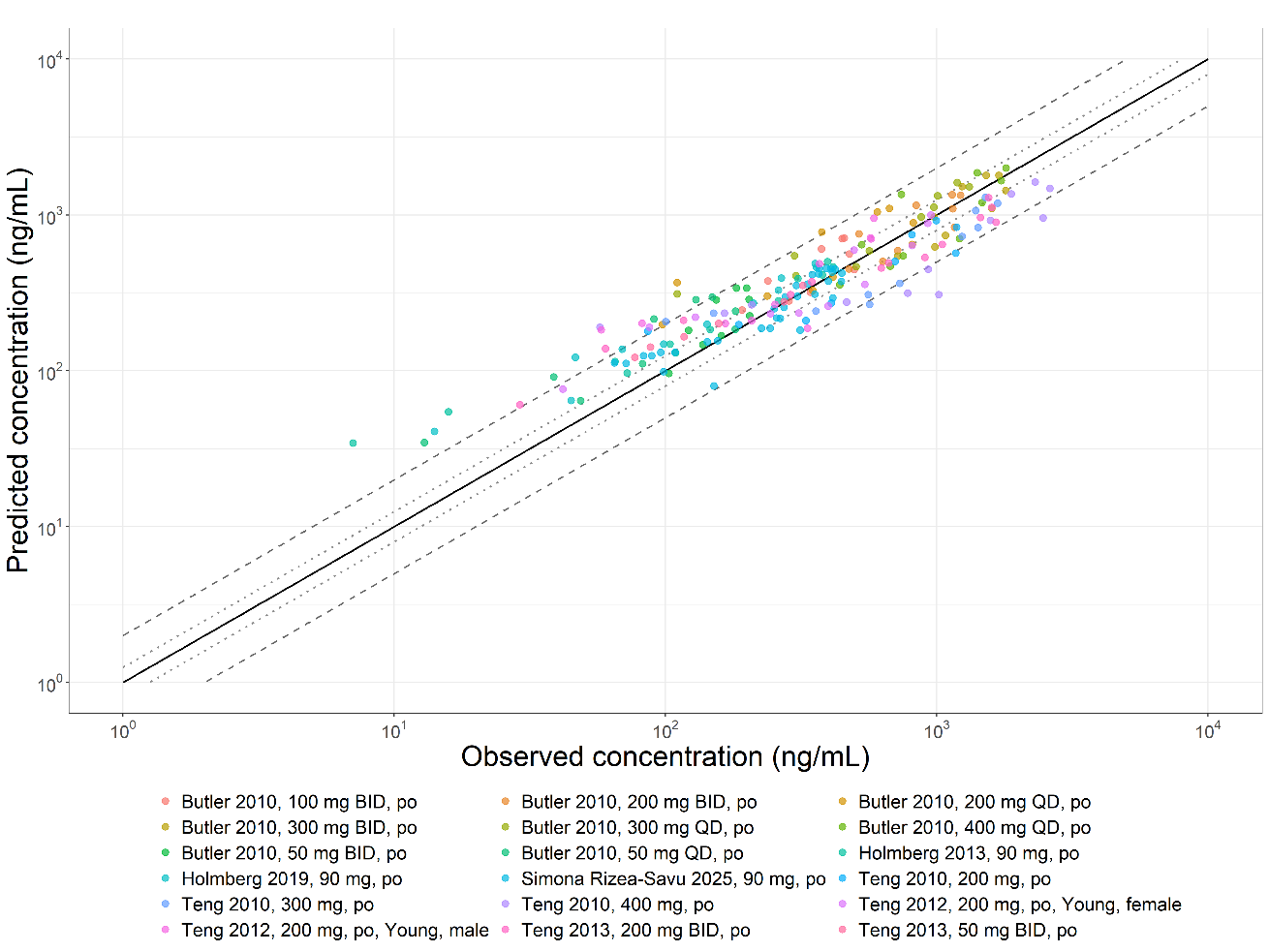 |
| FigS15: Predicted versus observed ticagrelor plasma concentration values for the training (a) and test (b) datasets in the base model. Solid lines represent the line of identity. Dotted lines indicate 1.25-fold deviation; dashed lines indicate 2-fold deviation. Details on administration protocols, study populations, and literature references are listed in table S10. | |

| A) | 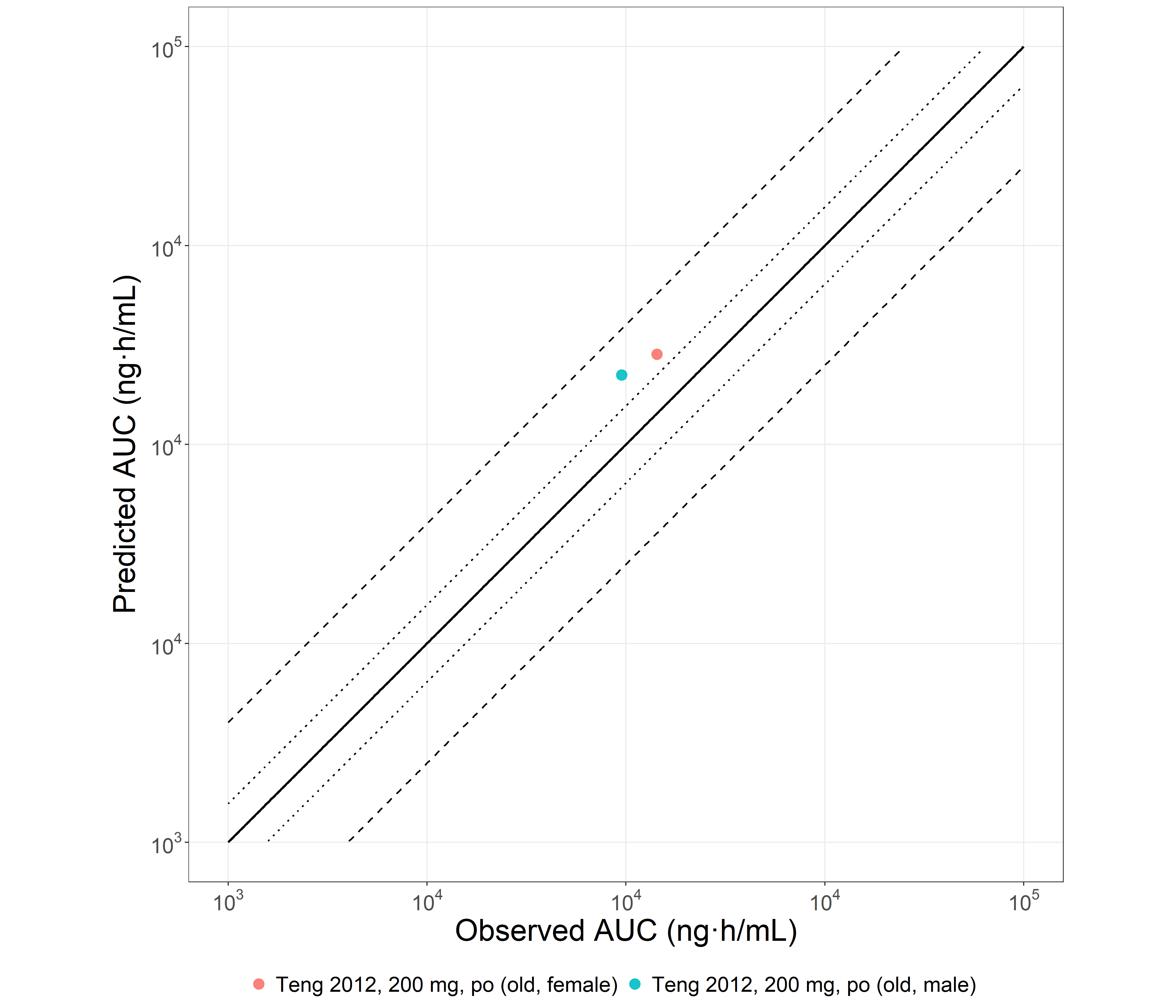 |
| --- | --- |
| B) | 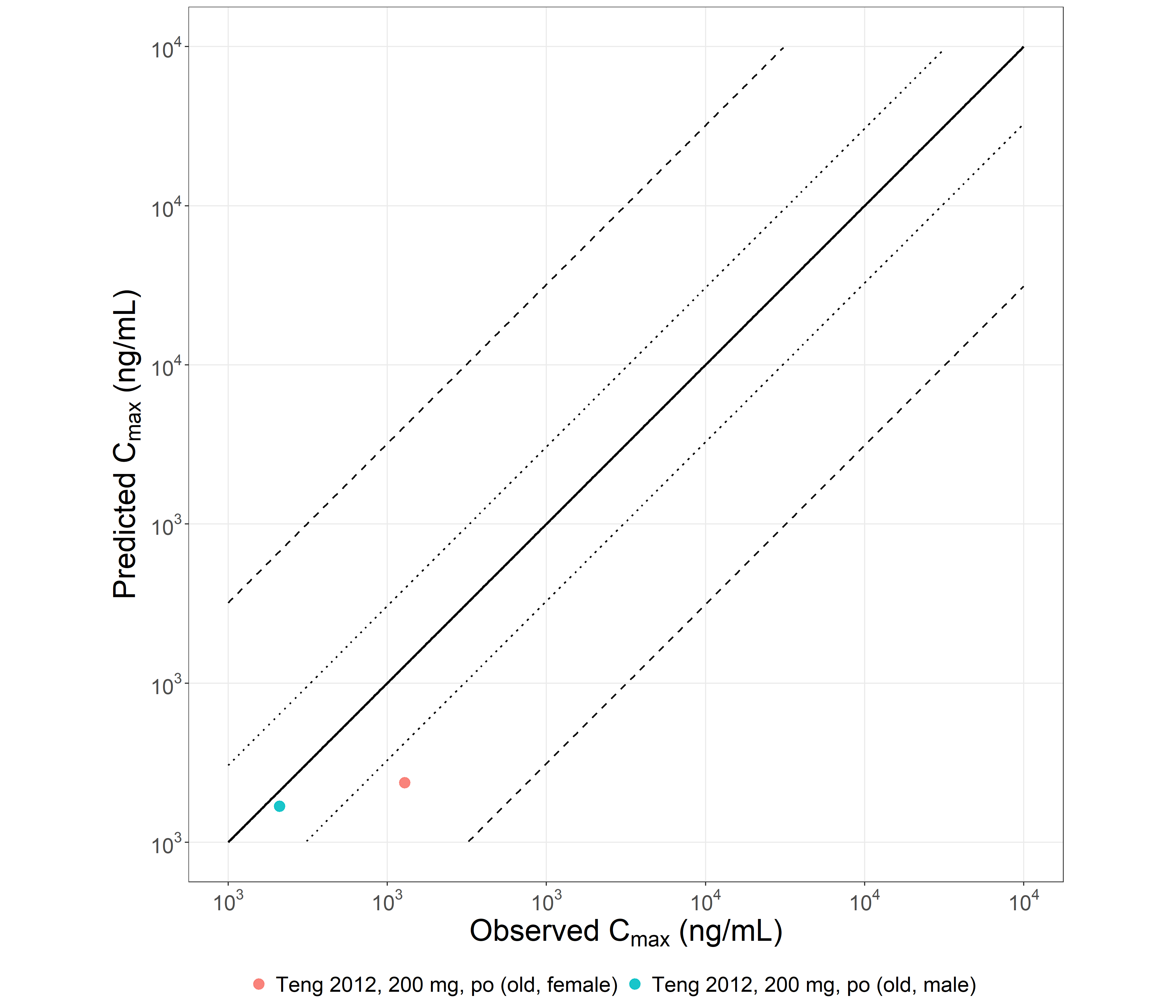 |
| C) | 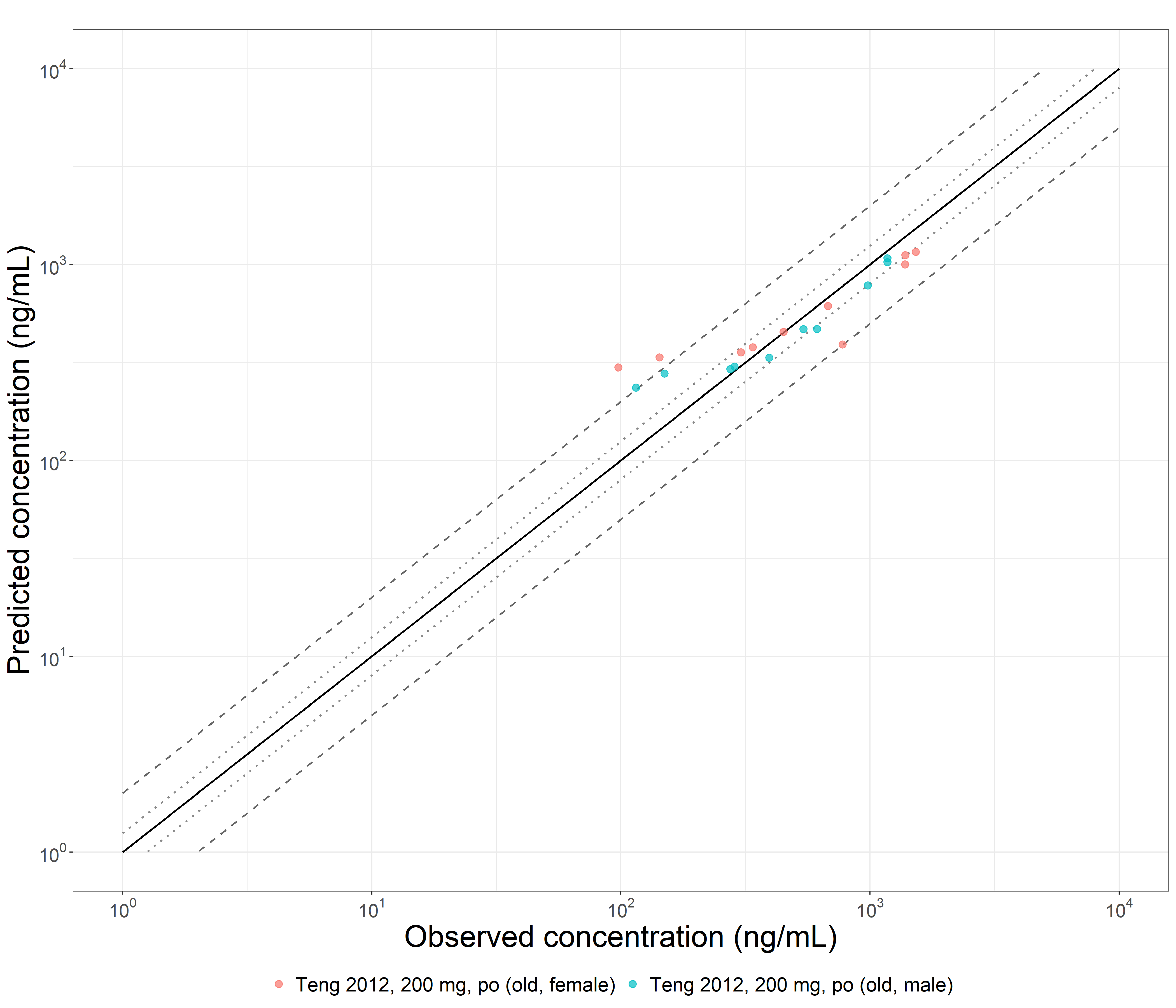 |
| FigS16: Predicted versus observed ticagrelor AUClast(a), Cmax (b) and plasma concentration (c) values for the geriatric population. Solid lines represent the line of identity. Dotted lines indicate 1.25-fold deviation; dashed lines indicate 2-fold deviation. Details on administration protocols, study populations, and literature references are listed in table S9. | |
